# Movement-responsive deep brain stimulation reinforces motor circuits in Parkinson’s disease

**DOI:** 10.64898/2026.08.20.26360021

**Authors:** Daryl J. Lawrence, Jiyeon Suh, Victoria Chang, Jeffrey A. Herron, Philip A. Starr, Simon J. Little

## Abstract

Deep brain stimulation is an established treatment for Parkinson’s disease but does not adapt to dynamic changes in brain state. Here, in four patients with sensing-enabled DBS systems, we evaluated a movement-responsive DBS (mDBS) paradigm that modulated subthalamic stimulation based on volitional motion decoded from cortical activity. During structured motor tasks, mDBS improved average forearm speed and mitigated the progressive bradykinetic slowing observed under constant-amplitude DBS (cDBS), accompanied by a cumulative increase in sensorimotor cortical beta activity and connectivity. In unconstrained, daily activities, mDBS lowered average bradykinesia severity and demonstrated progressive symptom reduction over hours of therapy, which gradually reversed upon switching to cDBS. These findings highlight the enhanced therapeutic benefit of mDBS and its potential to reinforce functional motor circuits in disorders of movement.

## Main Text

Many neurological and psychiatric disorders are characterized by severe network dysfunction that benefit from circuit-specific therapies, such as deep brain stimulation (DBS) (*1*, *2*). While conventional, constant-amplitude DBS (cDBS) is an effective treatment for advanced movement disorders, including Parkinson’s disease (PD), dystonia and essential tremor (*3–5*), it has several key limitations. First, it does not adapt to fluctuations in clinical state or dynamic brain activity associated with varying symptom severity, medication cycles, and voluntary movement (*6*). Second, while cDBS provides acute symptomatic relief, this effect can be rapidly reversed upon cessation of stimulation (*7–10*). Third, cDBS requires a compromise in stimulation amplitude to balance suppression of hypokinetic symptoms, such as bradykinesia (slowness of movement), rigidity and tremor, with the avoidance of hyperkinetic side effects, such as dyskinesia, often resulting in residual motor impairment (*11*, *12*).

Brain-computer interfaces (BCIs) have demonstrated accurate decoding of neural activity to facilitate control of external devices, such as cursors and prosthetic limbs (*13*, *14*). Recent advancements have highlighted the convergence of BCIs with neuromodulation to develop enhanced circuit-specific therapies (*6*, *15*). Specifically, closed-loop, adaptive DBS (aDBS) has demonstrated precise, real-time decoding of clinically-defined states, such as fluctuations in dopaminergic medication levels (*16*), or physiological biomarkers (*17*). Notably, subthalamic beta activity, which is associated with the rigid/akinetic state, has been used as an aDBS control signal leading to increased on-time (when symptoms were well-controlled) without troublesome dyskinesias compared to cDBS (*18*, *19*). Inspired by classic BCI-based decoding of motor activity, our group demonstrated a proof-of-principle of an alternative aDBS paradigm - movement-responsive DBS (mDBS) - which targeted volitional forearm motion during brief, constrained motor tasks in a single patient with PD (*20*). Results showed that sensorimotor cortical local field potentials (LFPs) could be used for precise decoding of forearm movement, leading to improved hand speeds during a keyboard typing task and a reduction in involuntary, dyskinetic movements during rest. Further, we recently validated the fidelity of movement decoding in unconstrained, naturalistic settings using cortical signals in a larger group cohort (*21*). However, to date the longer-term therapeutic effects of mDBS in PD, when implemented in real-world conditions, remain unexplored.

Recent studies suggest that movement-contingent stimulation of the basal ganglia may not only offer acute symptomatic relief but could also provide sustained motor improvements when stimulation is discontinued by leveraging neural plasticity (*9*, *22–24*). Evidence from mouse models demonstrates that pairing optogenetic stimulation of direct pathway striatal neurons (*23*) or striatal dopamine axons (*25*) with fast movements can result in cumulative and persistent increments in movement velocity and expression. Similar plasticity effects have been observed in patients with PD. Subthalamic DBS (STN DBS), administered exclusively during fast forearm movements in a tablet-based motor task, significantly improved movement speed in subsequent trials post-stimulation (*24*). While these effects suggested activity-dependent synaptic plasticity, they were transient - lasting only a few minutes - and observed under controlled, perioperative research conditions. Further evaluation of movement state-dependent stimulation is needed for clinical translation, to establish whether it can achieve therapeutic benefits over sustained periods in naturalistic environments, be integrated with ongoing therapy and strengthen functional motor circuits.

Here, we evaluated the mDBS approach chronically at home by conducting a blinded, randomized, crossover feasibility study with multiple repetitions involving four PD patients (eight independently-optimized hemispheres; Table S1). A data-driven pipeline was constructed to identify personalized sensorimotor cortical biomarkers of forearm movement. Using these biomarkers, we developed machine learning (ML) models capable of rapidly and accurately decoding motion. These models were embedded on implantable neurostimulators, enabling their use during both structured motor tasks and unconstrained, daily activities. Compared to constant-amplitude stimulation, mDBS improved forearm speeds during the motor tasks and reduced bradykinesia severity when administered over hours in unsupervised, naturalistic conditions. Beyond immediate symptomatic relief, we found evidence of progressive symptom improvement over time, consistent with activity-dependent plasticity mechanisms that could potentially be leveraged to drive restorative changes in motor control circuits (*23*).

### Participant information and study outline

Patients were recruited from the cohort of a previous study aimed at examining the physiological basis for PD-related motor symptoms (ClinicalTrials.gov: NCT03582891). Each participant had received bilateral implantation of the Summit RC+S system (Medtronic, Inc.) (*26*, *27*), which combined chronic neural recording with programmable adaptive stimulation (Fig. 1A). Quadripolar depth leads were surgically inserted into STN for stimulation and recording of LFPs (Fig. 1B). Four-contact electrocorticography (ECoG) arrays chronically positioned over the central sulcus enabled simultaneous, additional recording from the primary motor (M1) and somatosensory cortices (S1) (Fig. 1C).

**Fig. 1.**
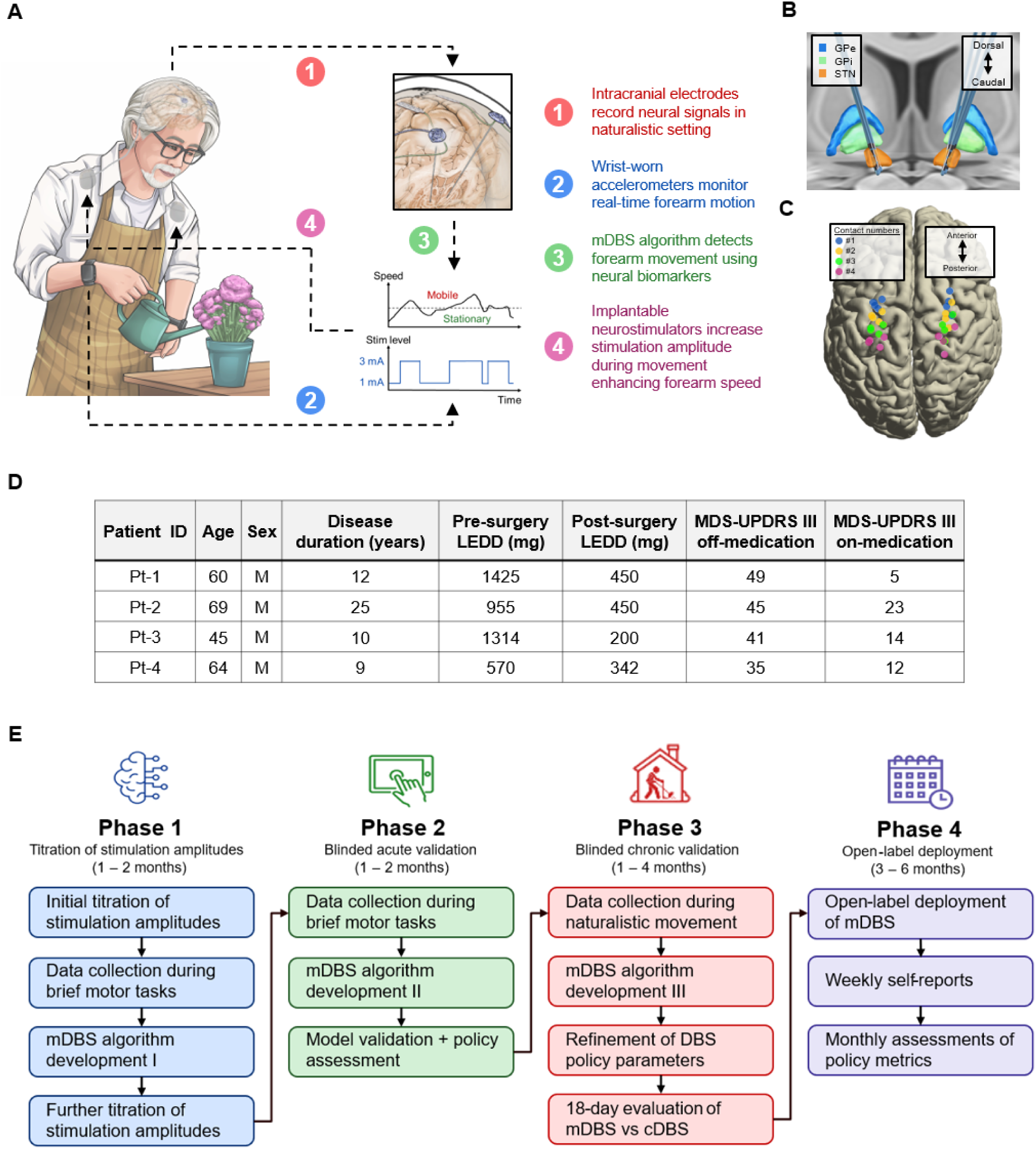
Movement-responsive DBS (mDBS) pipeline and patient characteristics. (A) Schematic of mDBS policy implemented during unconstrained, naturalistic motion. (B) Anatomic localization of implanted subcortical depth leads represented on a template brain in the globus pallidus externus (GPe), globus pallidus internus (GPi) and subthalamic nucleus (STN). (C) Cortical electrodes were also represented on a template brain. The four patients in our cohort used a non-overlapping, bipolar sensing configuration (Electrode pairs: 1-2 and 3-4). (D) Patient demographics and clinical characteristics, including age, sex, disease duration, pre-surgery and post-surgery levodopa equivalent daily dose (LEDD), as well as Movement Disorder Society Unified Parkinson’s Disease Rating Scale (MDS-UPDRS)-III off-medication and on-medication scores. (E) Workflow for the development, deployment and evaluation of mDBS paradigm.

We designed a four-phase protocol to systematically develop, validate, and deploy the mDBS system across increasing timescales and complexity (Fig. 1, D and E). Phase 1 involved the collection of neural and accelerometry signals during brief, structured motor tasks on a tablet computer to train the mDBS algorithm. Phase 2 comprised blinded, *acute* validation: the effect of mDBS on forearm movement speed was assessed during the same structured motor tasks. Phase 3 progressed to blinded, *chronic* validation: neural and accelerometry signals were collected in unconstrained, naturalistic settings to re-train the mDBS algorithm and investigate the effect of mDBS on bradykinesia and other PD-related symptoms. Phase 4 deployed mDBS continuously for 3-6 months in an open-label manner. All phases were conducted entirely remotely with patients at home, demonstrating real-world feasibility.

For each hemisphere, a movement disorder neurologist optimized stimulation parameters for cDBS, to ensure best-practice clinical care and a rigorous comparative control. Further, an optimal stimulation amplitude range for mDBS was established through iterative clinical assessment (Fig. 2A). This range was defined by a lower limit (the minimum amplitude required to mitigate breakthrough hypokinetic symptoms in the low dopaminergic state) and an upper limit (the maximum amplitude to effectively manage hypokinetic symptoms while avoiding DBS-related adverse effects, including dyskinesia and dysarthria, in the high dopaminergic state) during mDBS. The mDBS policy was designed to dynamically increase the stimulation amplitude from the lower limit to the upper limit during periods of detected movement.

**Fig. 2.**
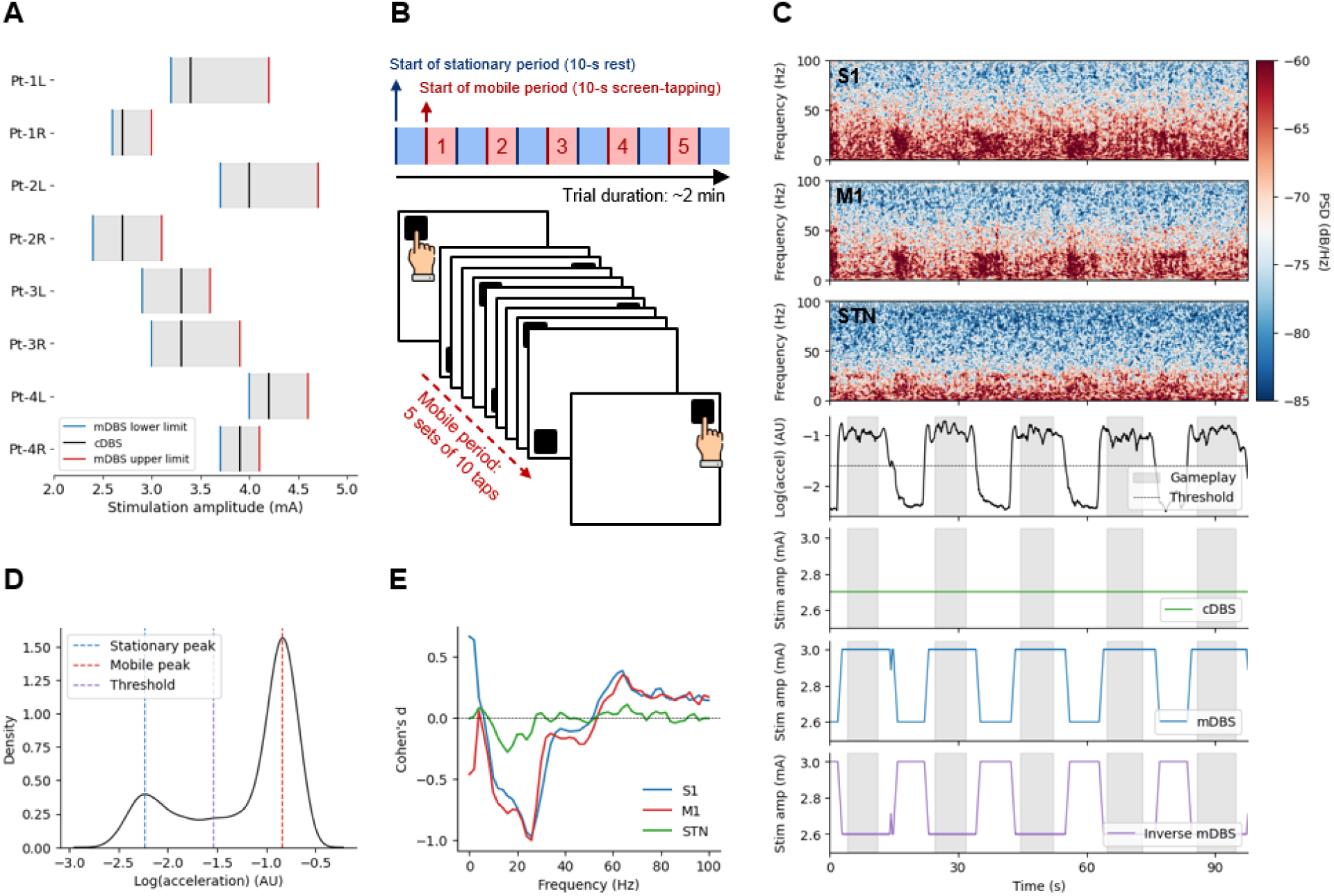
Overview of titrated stimulation amplitudes and single-hemisphere example of DBS policies administered during blinded, acute validation phase. (A) Optimal stimulation amplitude ranges for mDBS (upper and lower limits) and cDBS as determined by a movement disorders neurologist. (B) Schematic of the structured motor task – a tablet-based, screen-tapping game - used to assess forearm speeds and compare three stimulation policies: mDBS, cDBS, and inverse mDBS (stimulation amplitude was decreased during mobile periods and increased during stationary periods). Each game consisted of five sets of *stationary* (blue) and *mobile* (red) periods. (C) Single-hemisphere example of a spectrogram computed from LFPs recorded in S1, M1, and STN during the screen-tapping game. Simultaneously, forearm acceleration data were collected to distinguish between mobile and stationary states. Stimulation amplitudes shown illustrate the mechanism of each policy. (D) Accelerometry measurements from a single forearm during the structured motor tasks yielded a bimodal distribution, with peaks characterizing stationary and mobile periods. The average of these two peak acceleration values was used as the threshold to delineate between movement states. (E) Contralateral LFPs were used to compute power spectral densities (PSDs) (0–100 Hz, 500-ms epochs) for each brain region. Each PSD was assigned a binary movement state label (stationary or mobile) based on the accelerometry measurement at the end of the epoch. Cohen’s *d* effect sizes for PSDs in each region were calculated to evaluate their predictive power in classifying movement states. L and R indicate left and right forearms used by patients.

### Neural biomarkers of volitional movement

To identify biomarkers of movement for each patient and hemisphere, we first recorded neural and wrist accelerometry signals while patients performed a structured motor task (Fig. 2, B and C). This was a tablet-based game, which was conducted with their contralateral hand to the neural recordings whilst being stimulated at the high and low amplitudes identified for the mDBS policy. The game consisted of five sets, each containing a *stationary* period followed by a *mobile* period involving rapid screen-tapping. The wrist accelerometry measurements revealed a bimodal distribution of acceleration magnitudes, corresponding to ground-truth movement state labels of rest and movement (Fig. 2D and fig. S1A). We used the average of the two peaks in this distribution as the threshold to differentiate between stationary and mobile states. We then quantified differences in neural activity between these movement states using Cohen’s *d* effect sizes (*28*) to identify optimal cortical and subcortical kinematic biomarkers (Fig. 2E and Fig. 3, A and B). In both S1 and M1, significant movement-related desynchronization (MRD) was observed in the alpha (8-12 Hz) and beta (13-30 Hz) frequency bands, indicated by negative Cohen’s *d* effect sizes when comparing mobile versus stationary states (Fig. 3, C and D) (*29*). Concurrently, movement-related synchronization (MRS) was observed in the broadband gamma (60-100 Hz) range, as evidenced by positive Cohen’s *d* effect sizes reflecting higher spectral power during motion (*30*). Linear mixed models (LMMs) - with nested random effects for patient and hemisphere - revealed that while beta MRD was detectable within the STN (*β_0_* = 0.12 ± 0.06, *P* = 0.045) (*31*) its magnitude was significantly smaller than in cortical regions (S1: *β* = 0.49 ± 0.07, *P* < 10⁻^4^; M1: *β* = 0.47 ± 0.07, *P* < 10⁻^4^). Further, across the 0-100 Hz frequency range, mean absolute Cohen’s *d* values remained higher in S1 (*β* = 0.25 ± 0.03, *P* < 10⁻^4^) and M1 (*β* = 0.24 ± 0.03, *P* < 10⁻^4^) compared to the STN, demonstrating that cortical signals provided markedly higher discriminative power between movement states compared to subcortical signals (fig. S1B) (*21*, *32*).

**Fig. 3.**
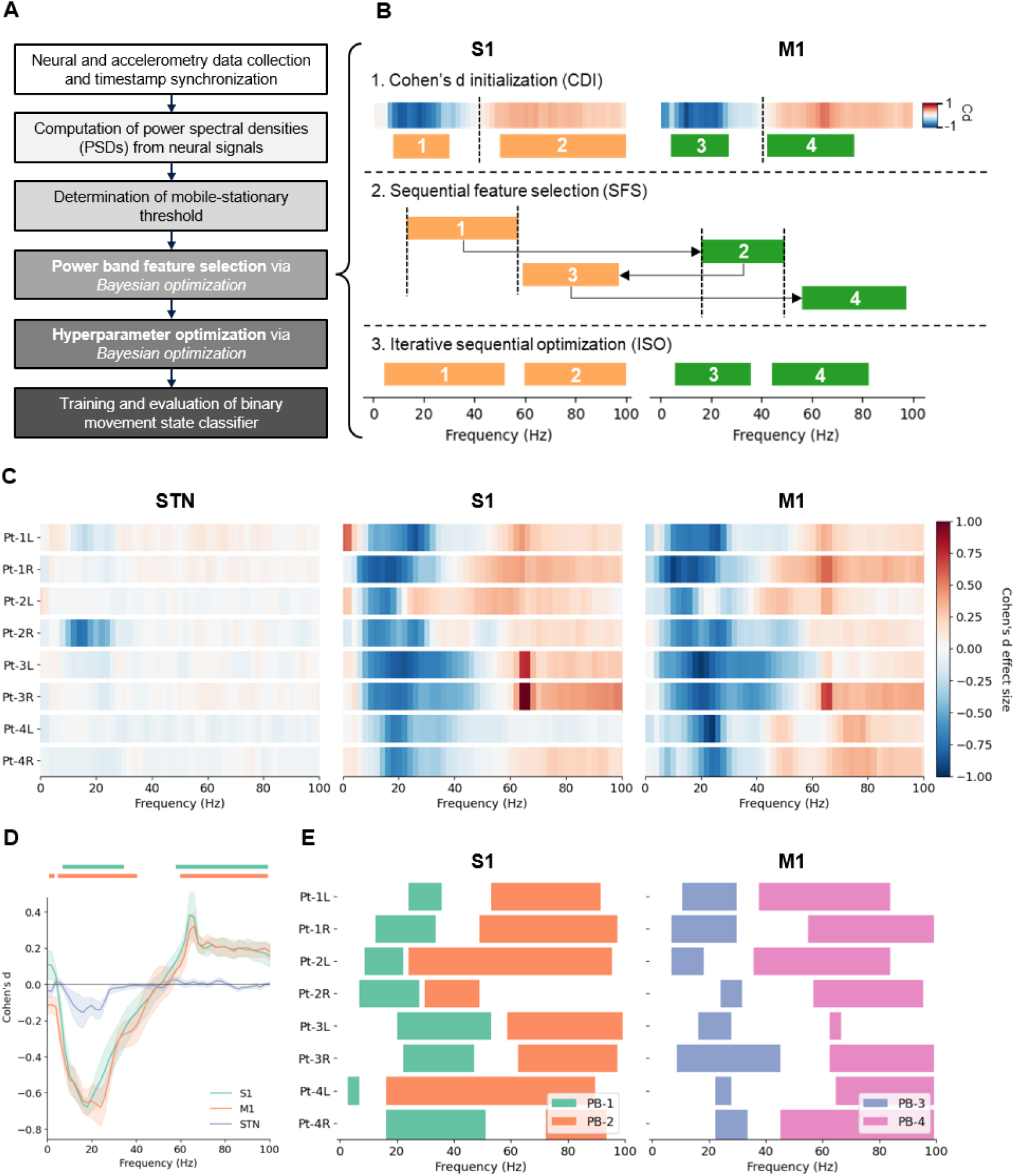
Overview of machine learning pipeline and selected power bands for blinded, acute validation. (A) Formalized procedure for developing the mDBS algorithm to decode movement states. (B) The Summit RC+S device supported four-feature linear models. To identify optimal power band (PB) features for each patient and hemisphere, the Cohen’s *d* initialization (CDI) algorithm was developed and compared to two other methods: sequential feature selection (SFS) and iterative sequential optimization (ISO). CDI leveraged Cohen’s *d* plots to identify frequency bands associated with movement-related desynchronization (MRD) and synchronization (MRS). These frequency bands were then used as constraints selecting PB features. SFS and ISO optimized PB features differently: SFS selected features sequentially while ISO optimized them simultaneously. Neither SFS nor ISO incorporated frequency constraints in their optimization processes. (C) Cohen’s *d* plots for each hemisphere and brain region. (D) Average Cohen’s *d* plots for each brain region. The horizontal bars above the plots indicate significant group-level movement biomarkers from each site after false-discovery rate correction (*P* < 0.05). (E) Selected cortical PBs yielding the highest five-fold cross-validation performance during feature selection across all algorithms. L and R indicate left and right forearms used by patients.

### Acute validation: movement-prediction during structured motor tasks

The Summit RC+S device can be embedded with a linear decoder that utilizes up to four spectral power bands (PBs) as weighted input features (*26*). Given the superior movement-discriminative power of the cortical signals (*20*, *21*, *32*), we prioritized S1 and M1 signals for feature selection (Fig. 3E). We then implemented a data-driven method, termed the Cohen’s *d* initialization (CDI) algorithm, to identify PBs for each patient and hemisphere. This approach utilized the MRD and MRS frequency ranges identified in the Cohen’s *d* plot for each hemisphere (Materials and Methods). To evaluate CDI, we compared it to two standard feature selection methods: sequential feature selection (SFS) and iterative sequential optimization (ISO) (*33*). Unlike CDI, SFS and ISO are exhaustive search methods that do not impose constraints on the frequency ranges for PB selection. All three methods used five-fold cross-validation to evaluate performance and employed Bayesian optimization to determine the frequency ranges of their respective PBs. ISO resulted in significantly lower balanced accuracy compared to CDI (*β* = -0.016 ± 0.007, *P* = 0.028). While CDI and SFS achieved similar model performance (Fig. 4A; *β* = -0.002 ± 0.007, *P* = 0.75), CDI required substantially less computation runtime (Fig. 4B; *β* = -28.4 ± 0.2 s, *P* < 10⁻^4^). These results demonstrate CDI’s ability to achieve a favorable tradeoff between model accuracy and computational efficiency when compared to SFS and ISO.

**Fig. 4.**
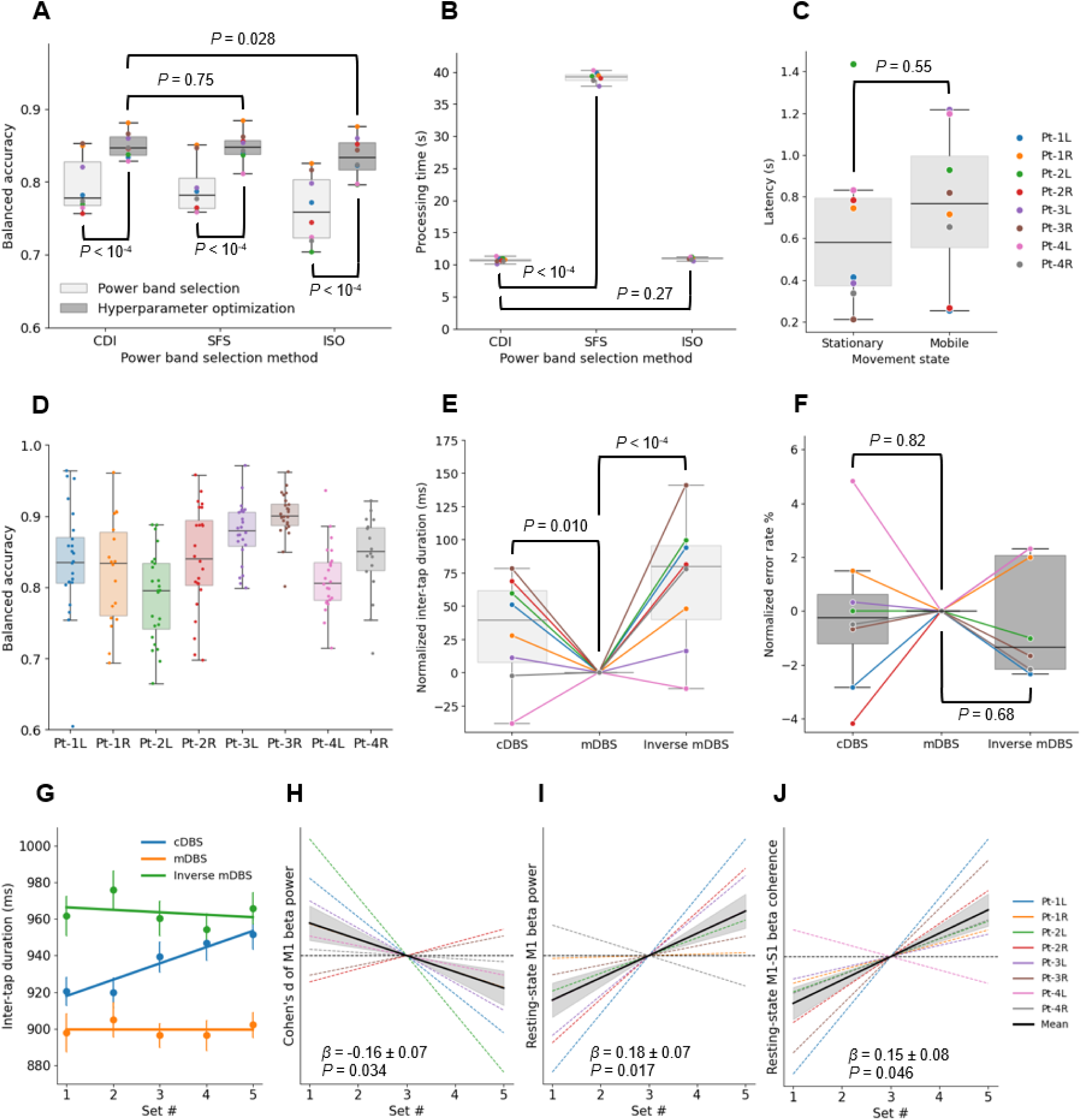
Performance and therapeutic effects of the mDBS algorithm during blinded, acute validation phase. (A) LMMs were used to compare the performances of the CDI, SFS and ISO feature selection algorithms (set as fixed effects). Mean balanced accuracy from five-fold cross-validation was computed for each hemisphere. Bayesian optimization was applied to tune smoothing hyperparameters for modeling forearm movement dynamics. PBs yielding the highest cross-validation performance were selected for the mDBS algorithm. (B) The processing times for each feature selection algorithm were also compared. (C) Study participants repeated the structured motor tasks to assess the performance of the mDBS algorithm and compare the effects of each DBS policy – mDBS, cDBS and inverse mDBS – on forearm speed. The average latency for the detection of stationary and mobile states was computed by determining the amount of time the predicted movement states lagged (positive latency values) or led (negative latency values) the ground truth movement states. (D) The balanced accuracy of the mDBS algorithm was also calculated. (E) LMMs were used to analyze the time between consecutive screen taps, termed the inter-tap duration, under each DBS policy (set as fixed effects). For more succinct visualization of the data, normalized inter-tap duration was plotted by subtracting the mean inter-tap duration under mDBS for each patient and hemisphere. (F) LMMs were also used to compare the screen-tapping errors caused by overshoots or undershoots during the motor tasks under each DBS policy (set as fixed effects). Normalized error rates were calculated by subtracting each patient’s mean error rate under mDBS and plotted. (G) The regression lines for the mean inter-tap duration (across patients and hemispheres) for each of the five sets during the screen-tapping game under each DBS policy was plotted for easy visualization of the cumulative changes in forearm speed. Pearson *r* correlation coefficients modeled the variation in (H) median resting-state M1 beta power (during only stationary periods), (I) Cohen’s *d* effect size of M1 beta power and (J) M1-S1 beta coherence over the five sets of the screen-tapping game under mDBS. In (H - J), the average regression lines for each patient and hemisphere, as well as the group-level average regression line (solid black), were normalized by subtracting the r coefficients for cDBS from those for mDBS. The standard error of the mean (grey shading) was also included. Statistical comparisons were performed using LMMs on the r coefficients (without normalization) for cDBS and mDBS. Patients and hemispheres were included as random effects in each LMM. L and R indicate left and right forearms used by patients.

At the group-level, the selected PBs were centered in the beta band (S1: 25 ± 4 Hz; M1: 23 ± 2 Hz) or the gamma band (S1: 68 ± 5 Hz; M1: 73 ± 3 Hz). PBs centered in the gamma band were significantly wider than those centered in the beta bands (S1: *P* = 0.013; M1: *P* = 0.008), consistent with broadband gamma synchronization during movement (*34*). Incorporating temporal smoothing hyperparameters to accommodate individual movement dynamics significantly improved classification performance for all three algorithms (*P* < 10⁻^4^) (*35*).

### Acute validation: mDBS improves forearm movement speed during motor tasks

We evaluated the test-time performance of the mDBS algorithm by comparing predicted movement states against concurrent accelerometry during gameplay on the tablet computer. The algorithm detected state transitions with a mean latency (± SEM) of 0.7 ± 0.1 s, ensuring patients received appropriate stimulation for the majority of each mobile and stationary period (Fig. 4C; Mean duration of each period ± SEM: 10.8 ± 0.4 s), and achieved a mean balanced accuracy of 0.84 ± 0.01, matching the offline cross-validation performance of 0.850 ± 0.006 (Fig. 4D; β = -0.012 ± 0.009, *P* = 0.24). To assess therapeutic impact, we measured motor performance during the screen-tapping game under three conditions: mDBS (stimulation amplitude increased during movement), cDBS (constant-amplitude stimulation), and inverse mDBS (stimulation amplitude decreased during movement). The inverse mDBS control condition was included to rule out the possibility that any benefit of mDBS might be solely attributable to the intermittency of stimulation rather than its movement state-specific modulation. mDBS significantly reduced inter-tap durations compared to both inverse mDBS (*β* = 68 ± 12 ms, *P* < 10⁻^4^) and cDBS (*β* = 32 ± 12 ms, *P* = 0.010), reflecting improved forearm movement speeds (Fig. 4E). Notably, this improvement in forearm speed occurred without increasing error rates (inverse mDBS: *β* = -0.3 ± 0.8 %, *P* = 0.68; cDBS: *β* = -0.2 ± 0.8 %, *P* = 0.82; Materials and Methods), indicating that tapping accuracy was equivalent across all three conditions (Fig. 4F). These results demonstrate that selectively increasing stimulation amplitude during movement enhances motor speed without triggering troublesome dyskinetic movements that could compromise precision.

### Acute validation: mDBS modulates progression of forearm speed and cortical beta MRD

To assess potential changes in forearm speed during the screen-tapping game, we calculated the median inter-tap duration for each of the five sets of stationary-mobile periods, followed by the Pearson *r* correlation coefficient across these sets. Our findings revealed a significant group-level increase in inter-tap duration under cDBS, indicating a cumulative bradykinetic slowing of forearm speed (Fig. 4G; *β*_0_ = 0.24 ± 0.08, *P* = 0.029). Compared to cDBS, there was no significant group-level change in forearm speed across the five sets under mDBS. This suggests that movement state-contingent stimulation may have reinforced functional motor circuits, resulting in an antibradykinetic effect that mitigated the slowing observed with cDBS (fig. S2A; *β* = -0.24 ± 0.07, *P* = 0.001). Similarly, no significant change in forearm speed over the five sets was found under inverse mDBS (fig. S3A; *β* = -0.25 ± 0.07, *P* = 0.001). However, this may have been attributable to a ceiling effect, as the inverse DBS condition showed the highest average inter-tap durations compared to both cDBS (*β* = 35 ± 13 ms, *P* = 0.005) or mDBS (*β* = 68 ± 12 ms, *P* < 10⁻^4^).

We also observed electrophysiological biomarkers consistent with neural reinforcement induced by mDBS during the motor tasks. Specifically, under mDBS compared to cDBS, there was a cumulative increase in M1 beta MRD across the five sets of the screen-tapping game, as evidenced by a reduction in Cohen’s *d* effect size (i.e., more negative Cohen’s *d* value; Fig. 4H; *β* = -0.16 ± 0.07, *P* = 0.034). This result was driven by a progressive increase in the median resting-state M1 beta power over the five *stationary* periods (Fig. 4I; *β* = 0.18 ± 0.07, *P* = 0.017). In contrast, over the five *mobile* periods, there was no progressive change in M1 beta power (fig. S2B; *β* = -0.03 ± 0.08, *P* = 0.67). Additionally, mDBS was associated with a cumulative increase in resting-state M1-S1 beta coherence (over the five *stationary* periods) under mDBS compared to cDBS (Fig. 4J; *β* = 0.15 ± 0.08, *P* = 0.046). Over the five *mobile* periods, there was no progressive change in M1-S1 beta coherence (fig. S2C; *β* = 0.02 ± 0.08, *P* = 0.81) under mDBS compared to cDBS. Under the inverse mDBS condition, no significant change was detected in M1 beta MRD (fig. S3B; *β* = -0.12 ± 0.07, *P* = 0.10), median M1 beta power (fig. S3, C and D; Resting state: *β* = 0.08 ± 0.07, *P* = 0.33; Mobile state: *β* = -0.09 ± 0.08, *P* = 0.23) or M1-S1 beta coherence (fig. S3, E and F; Resting state: *β* = 0.02 ± 0.08, *P* = 0.76; Mobile state: *β* = -0.10 ± 0.08, *P* = 0.21) over the five stationary or mobile periods.

### Chronic validation: movement prediction in unconstrained, naturalistic settings

Extending mDBS from brief, structured motor tasks to unsupervised daily activities required validating the stability of the movement-predictive algorithm across diverse, unrestricted movements. The inverse mDBS control policy was excluded from the chronic validation phase because it resulted in reduced forearm speeds during the structured motor tasks compared to mDBS or cDBS. The mDBS algorithm was re-trained using neural and wrist accelerometry data collected while patients performed everyday activities at home (∼6 hours recorded; Materials and Methods). Analysis of these measurements revealed that the neural biomarkers were consistent with those found during the structured motor tasks (Fig. 5, A, B and C). Specifically, cortical alpha-beta MRD and broadband gamma MRS remained the dominant biomarkers of movement (*29–31*). However, absolute Cohen’s *d* effect sizes decreased compared to structured motor tasks (fig. S4A; *P* < 0.008), reflecting the greater complexity and heterogeneity of naturalistic behavior (*36*). Through feature selection, CDI was found to outperform ISO albeit at the significance threshold (fig. S4B; *β* = -0.020 ± 0.009, *P* = 0.050) and required less computation runtime than SFS (fig. S4C; *β* = -256 ± 11 s, *P* < 10⁻^4^). This demonstrated that the frequency constraints used in CDI to reduce the power band search space improved the efficiency of the feature selection process while preserving model performance (Fig. 5D). Movement state-decoding performance was further enhanced by tuning the temporal smoothing hyperparameters of the mDBS algorithm to model individual behavioral patterns, consistent with the findings in the acute validation phase.

**Fig. 5.**
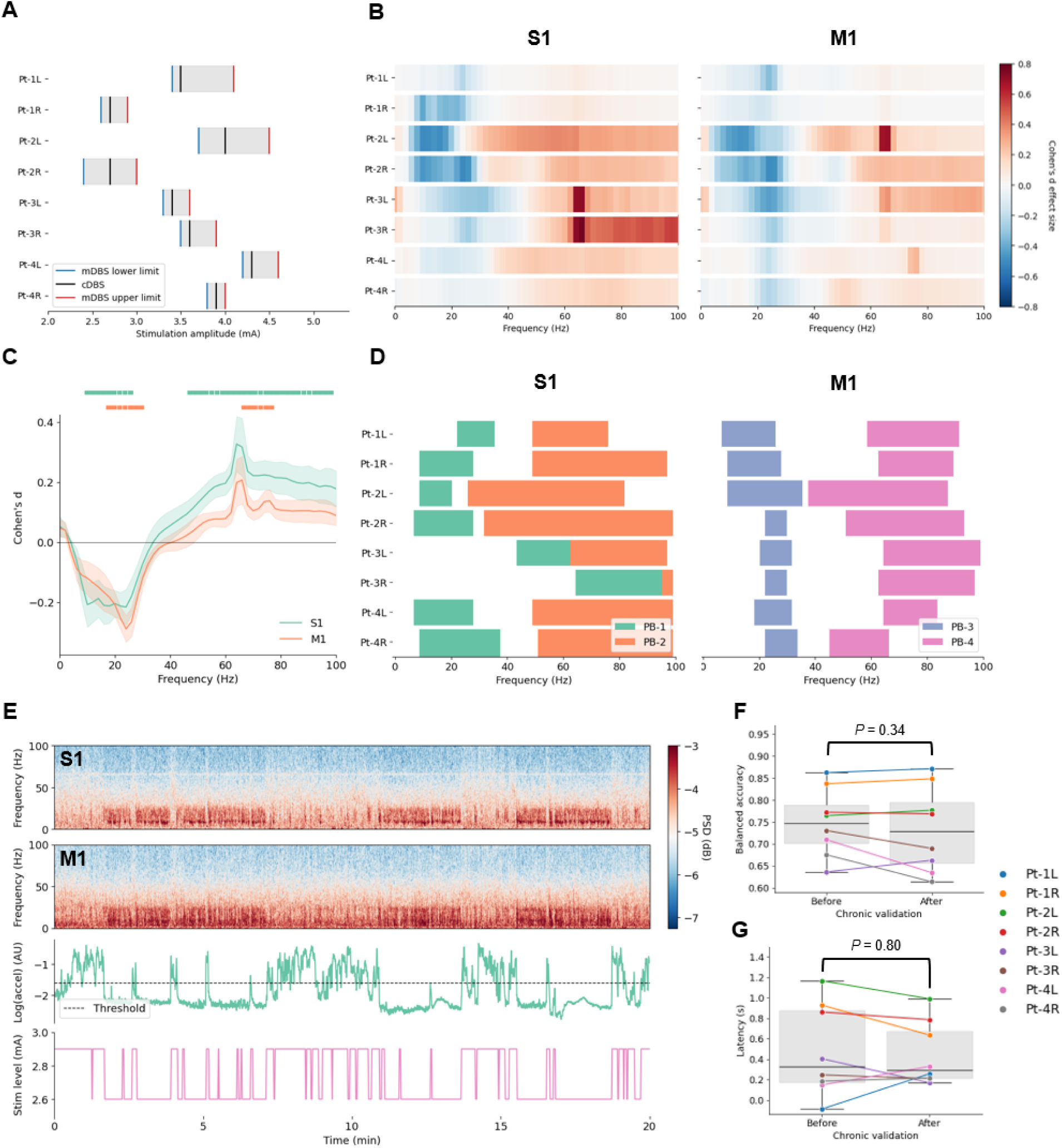
Development and performance of mDBS during the blinded, chronic validation phase. (A) Stimulation amplitude ranges for mDBS (upper and lower limits) and cDBS refined prior to the 18-day evaluation period. (B) Cortical Cohen’s *d* plots for individual hemispheres. (C) Average Cohen’s *d* plots for S1 and M1. The color-coded horizontal bars above the plots indicate significant group-level biomarkers of naturalistic movement after false-discovery rate correction (*P* < 0.05). (D) Optimal power bands selected for decoding naturalistic movement states. (E) Single-hemisphere example of a spectrogram computed from local field potentials (LFPs) recorded in S1 and M1 during unconstrained, unsupervised movement while patients were at home. Simultaneously, accelerometry data were collected to distinguish between mobile and stationary states and used to re-train the mDBS algorithm for the chronic validation phase. The mDBS algorithm triggered an increase in stimulation amplitude whenever movement was detected using the selected cortical biomarkers. Neural and accelerometry data were also recorded over a single day *before* and *after* the evaluation period and the average (F) balanced accuracy and (G) latency during these sessions were computed to evaluate the performance of the mDBS algorithm. Latency was determined by computing the average amount of time the predicted movement states lagged the ground truth movement states. Group-level comparisons in (F) & (G) were performed using LMMs with patients and hemispheres included as random effects. L and R indicate left and right forearms used by patients.

### Chronic validation: mDBS reduces bradykinesia severity in naturalistic settings

To ensure that stimulation amplitudes titrated during the acute validation phase remained optimal during the chronic validation phase, mDBS and cDBS were administered over several days and the stimulation amplitudes were refined based on patient feedback and motor assessments using Parkinson’s KinetiGraph (PKG) watches (*37–39*). The clinical effects of mDBS and cDBS were then evaluated over an 18-day period using a counter-balanced design with multiple repetitions (a total of 36 half-day sessions per subject where a single DBS policy was administered during each half-day session while patients were awake; Fig. 5E). Neural and accelerometry signals were recorded before and after this evaluation period to assess the mDBS algorithm’s performance. Before the evaluation period, the mean balanced accuracy of the mDBS algorithm was calculated as 0.75 ± 0.03 (Fig. 5F). While this was lower than in the acute validation phase, it was expected given the lower signal-to-noise ratio in naturalistic behavior. The mean latency was computed as 0.48 ± 0.15 s (Fig. 5G), indicating the algorithm’s responsiveness to changes in movement state. Importantly, no significant change in balanced accuracy (*β* = 0.02 ± 0.02, *P* = 0.33) or latency (*β* = 0.03 ± 0.13, *P* = 0.80) was observed over the 18-day evaluation period, demonstrating the stability of the mDBS algorithm’s performance.

Objective motor assessments from PKG watches showed a significant group-level reduction in bradykinesia under mDBS compared to cDBS (Fig. 6A; *β* = -1.9 ± 0.5 PKG units, *P* < 10⁻⁴), without a significant change in dyskinesia (Fig. 6B; *β* = 0.14 ± 0.10 PKG units, *P* = 0.15).

**Fig. 6.**
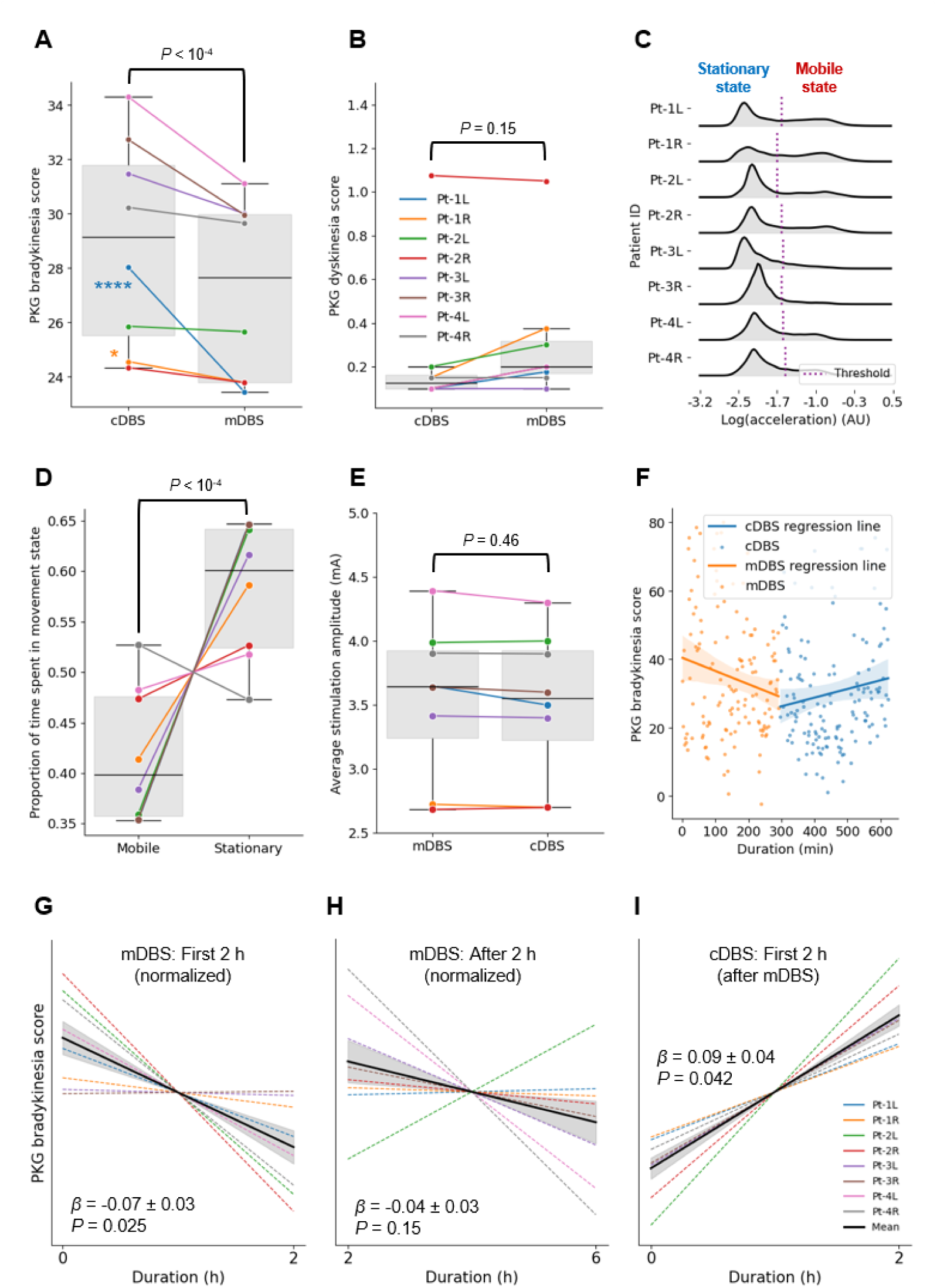
Therapeutic effects of mDBS during blinded, chronic validation phase. Median (A) bradykinesia and (B) dyskinesia severity scores, measured using PKG watches, were plotted for each patient and hemisphere under mDBS and cDBS conditions. LMMs quantified the differences between DBS policies. (C) Accelerometry measurements from the contralateral forearm, recorded via Apple watches, were used to re-train the mDBS algorithm for the chronic validation phase. The thresholds for stationary and mobile states were determined during the acute validation phase. (D) The average proportion of time spent in each movement state (stationary vs mobile) under mDBS during the 18-day evaluation period was computed from stimulation log files and quantified using LMMs. (E) The average stimulation amplitude under mDBS and cDBS was also computed from stimulation log files and quantified using LMMs. (F) A single-hemisphere, single-session example of PKG bradykinesia scores is shown for blinded administration of mDBS (∼6 hours) followed by cDBS (∼6 hours). Pearson *r* correlation coefficients were used to model the variation in PKG bradykinesia scores during two time periods: (G) the first 2 hours of each DBS policy and (H) the remaining ∼4 hours. For each patient and hemisphere, the average regression line was plotted, along with the group-level average regression line (solid black). To enhance visualization, the data were normalized by subtracting the r coefficient for cDBS from that for mDBS. (I) Pearson *r* correlation coefficients were also used to model the variation in PKG bradykinesia scores during the first 2 hours of cDBS after switching from mDBS. These values were normalized by subtracting the r coefficients from the first 2 hours of cDBS when it was administered prior to mDBS. The standard error of the mean was shown as grey shading for the average regression lines in (G - I). Statistical comparisons were performed using LMMs on the r coefficients (without normalization) for cDBS and mDBS. Patients and hemispheres were included as random effects in each LMM. L and R indicate left and right forearms used by patients.

Individual analyses revealed that Pt-1, who had the most severe baseline off-medication bradykinesia and residual fluctuations in motor function (indicated by the largest difference in on- and off-medication MDS-UPDRS III scores, as shown in Fig. 1D) experienced significant bilateral improvements in PKG bradykinesia scores (*P* < 0.018). Excluding Pt-1, the group-level reduction in PKG bradykinesia scores remained significant for the remaining patients (*β* = -1.6 ± 0.6 PKG units, *P* = 0.010), though within-subject analyses were non-significant (*P* > 0.05). Additionally, no significant changes were observed in PKG tremor scores (*β* = 0.3 ± 0.3 PKG units, *P* = 0.18) when comparing mDBS to cDBS. Self-reported scores of dystonia, gait, or dysarthria severity (fig. S5, A, B and C; FDR-corrected *P* > 0.17) - which were not captured by the PKG watches - also showed no significant difference between DBS policies. These findings collectively indicate a group-level reduction in bradykinesia under mDBS compared to cDBS without an increase in opposite motor side effects (dyskinesias) typically observed with excessive constant-amplitude approaches.

To confirm that the group-level reduction in bradykinesia under mDBS was not simply caused by an increase in average stimulation amplitude, we analyzed the stimulation log files (Materials and Methods). When mDBS was administered during the evaluation period, the proportion of time spent in the mobile state (higher stimulation amplitude) was significantly lower than the time spent in the stationary state (lower stimulation amplitude) (Fig. 6, C and D; *β* = -23.1 ± 1.5 %, *P* < 10^-4^). Consequently, we found no significant difference in average stimulation amplitude between mDBS and cDBS (Fig. 6E; *β* = 0.03 ± 0.04 mA, *P* = 0.46). These findings support that the group-level reduction in bradykinesia under mDBS was due to the selective provision of additional stimulation during volitional movement, rather than simply an overall increase in average stimulation amplitude.

### Chronic validation: cumulative therapeutic enhancement over hours

During the 18-day evaluation period, PKG measurements were used to analyze the temporal dynamics of bradykinesia severity under both mDBS and cDBS (Materials and Methods). A single-hemisphere example illustrating the variation in PKG bradykinesia scores over the course of a single session, with a switch midway from mDBS to cDBS, is presented in Figure 6F. Specifically, we sought to investigate whether mDBS might drive progressive accumulation in symptom benefit compared to amplitude-matched cDBS. We found that mDBS resulted in a cumulative group-level reduction in PKG bradykinesia scores over the initial two hours compared to cDBS (Fig. 6G; *β* = -0.07 ± 0.03, *P* = 0.025). There was no statistically significant change in PKG bradykinesia score over the following four hours (Fig. 6H; *β* = -0.04 ± 0.03, *P* = 0.15). Further, switching from mDBS to cDBS after ∼6-hours caused a significant group-level gradual rebound (increase) in bradykinesia severity (Fig. 6I; *β* = 0.09 ± 0.04, *P* = 0.042). This finding suggests that the therapeutic effects of mDBS may engage a form of activity-dependent plasticity that gradually reverses under constant-amplitude stimulation, which lacks movement-specific targeting (*24*, *25*). Importantly, analysis of the stimulation log files confirmed that the progressive reduction in bradykinesia observed under mDBS was not attributable to differences in average stimulation amplitude compared to cDBS (*P* = 0.46).

### Long-term deployment reveals therapeutic stability

During the follow-on, open-label deployment of mDBS (Phase 4: 3 - 6 months, continuous 24/7 operation), patient self-reports indicated a slight reduction in bradykinesia severity at the group-level (*β* = -0.61 ± 0.30 %, *P* = 0.047). Individual analyses revealed that Pt-1 experienced a further progressive decrease in self-reported bradykinesia severity (*P* = 0.005) during the open-label deployment phase. The other patients in the cohort reported stable therapeutic benefits without worsening of bradykinesia symptoms (*P* > 0.09). Additionally, there was no evidence of increased dyskinesia severity (*β* = -0.12 ± 0.20 %, *P* = 0.55) or changes in sleep quality (*β* = -0.2 ± 0.4 %, *P* = 0.62) under mDBS. Algorithm stability analysis during awake hours showed that patients spent less time in the mobile state compared to the stationary state (fig. S5D; *β* = -27 ± 3 %, *P* < 10⁻⁴), consistent with the chronic validation phase. Additionally, there was no significant change in the proportion of time spent in each movement state (during awake hours) over the open-label deployment phase (fig. S5E; *β* = -0.3 ± 1.1 %, *P* = 0.78), demonstrating no evidence of classifier drift. These findings reaffirm the stability in the algorithm performance and therapeutic efficacy of mDBS during extended use.

## Discussion

We developed and validated a movement-responsive DBS paradigm for chronic, naturalistic deployment in Parkinson’s disease. The mDBS algorithm dynamically modulated stimulation amplitudes in real-time according to movement states decoded from cortical signals. The algorithm demonstrated robust precision and stability across multiple timescales - from milliseconds (during blinded, acute validation) to months (during open-label deployment) - and across contexts, including structured motor tasks and unconstrained daily activities. During the motor tasks, mDBS led to improved forearm speeds compared to conventional, constant-amplitude stimulation. mDBS also overcame the progressive bradykinetic slowing observed under cDBS, suggesting that it reinforces functional motor circuits. This effect was accompanied by a cumulative increase in M1 beta MRD, resting-state M1 beta power, and resting-state M1-S1 beta coherence during the motor tasks, identified as potential biomarkers of neural reinforcement. In naturalistic settings, mDBS reduced group-level bradykinesia severity compared to cDBS. Further, the progressive reduction in symptoms observed over several hours under mDBS, and its reversal upon switching to cDBS, suggests that mDBS may leverage activity-dependent plasticity mechanisms that could drive cumulative motor improvements with prolonged use. This capability could be exploited to selectively reinforce functional motor circuits in PD and other movement disorders.

### Mechanistic insights on cortico-basal motor control

The mDBS paradigm is grounded in the gating theory of basal ganglia function, which posits that in PD, excessive inhibitory output from the basal ganglia to thalamo-cortical circuits impedes the initiation and execution of volitional motor plans, resulting in bradykinesia (*40–42*). DBS alleviates bradykinesia by lowering the barrier to movement and disinhibiting the execution of these motor plans (*43*). However, constant-amplitude stimulation represents a trade-off between minimizing both hypokinetic symptoms and hyperkinetic side effects (*6*). The fixed-amplitude approach may provide insufficient stimulation to disinhibit voluntary motor plans leading to persistent bradykinesia and rigidity. Conversely, it may inadvertently disinhibit involuntary motor plans, causing dyskinetic movements. Movement-responsive DBS overcomes these limitations by increasing the stimulation amplitude when volitional motion is detected, thereby selectively disinhibiting volitional motor programs.

### Complementary insights from acute and chronic validation phases

The acute and chronic validation phases of the study provided complementary evidence regarding the mechanism and therapeutic efficacy of mDBS. During the acute validation phase, forearm speeds were significantly lower under the inverse mDBS policy - wherein the stimulation amplitude was decreased, rather than increased, upon the detection of volitional forearm motion - compared to cDBS or mDBS (*20*). Consistent with the basal ganglia gating theory, this reduction in motor performance suggests that the decreased stimulation during movement likely impeded the disinhibition of voluntary motor plans, thereby exacerbating bradykinesia (*43*). These results support that the therapeutic benefits of mDBS are dependent on selectively increasing stimulation amplitude during neural states associated with movement.

Further, our chronic validation of the mDBS system in naturalistic settings allowed us to assess its therapeutic efficacy under real-world conditions over an extended 18-day period, using multiple within-subject trial blocks. We observed a significant group-level reduction in bradykinesia severity with mDBS compared to cDBS without a corresponding increase in opposite motor side effects, such as dyskinesias. However, the study cohort of four patients showed inter-participant variability. The patient with the most severe off-medication bradykinesia severity and residual motor fluctuations experienced the greatest therapeutic benefit from mDBS. While the remaining three patients still yielded a significant group-level reduction in PKG bradykinesia scores, they did not demonstrate statistically-significant motor improvements at the individual level. Consequently, the small, all-male sample of our study limits the generalizability of these results across the heterogeneous PD population, underscoring the need for larger studies. Nevertheless, our findings highlight the potential value of stratified enrollment in future studies based on baseline off-medication bradykinesia severity and persistent symptom fluctuations to optimize candidate selection for mDBS treatment.

### Activity-dependent plasticity: implications for therapeutic mechanisms

Therapeutic interventions for PD need not act on nigrostriatal cell loss to alter the trajectory of motor disability. Paradoxical kinesia, together with the finding that patients with PD can produce fast, accurate movements when cued or incentivized but require far more attempts to do so otherwise, indicates that bradykinesia reflects in part an altered implicit valuation of movement vigor rather than lost motor capacity (*44*, *45*). Chronic dopamine depletion biases the cost-benefit computation that sets vigor (*46*), entrenching a slower default and amplifying a focal cellular deficit into a distributed behavioral one. Movement-responsive stimulation likely acts on this downstream layer. Because detection requires volitional movement to exceed a decoder threshold, stimulation amplitude is increased preferentially during a patient’s more vigorous movements, selectively reinforcing the faster part of the distribution. Speed-contingent basal ganglia stimulation has been shown to produce cumulative, persistent increments in velocity in rodents (*23*), and an analogous process in humans, triggered using neural activity, could raise learned vigor by reinforcing functional motor circuits without impacting dopaminergic cell survival.

In the acute validation phase, progressive slowing of forearm speed during the motor task was observed under cDBS. However, this was absent under mDBS, suggesting a potential cumulative antibradykinetic effect induced by motor circuit reinforcement. We identified potential electrophysiological biomarkers of this neural reinforcement effect. Specifically, under mDBS compared to cDBS, there was a cumulative increase in movement-related beta desynchronization in the motor cortex, driven by an increase in resting-state beta power. These findings align with those of Cavallo et al., who reported, in a single subject, that brief, state-dependent subthalamic stimulation acutely enhanced post-movement beta rebound compared to unstimulated movements (*24*). Increased cortical beta MRD has also been linked to the selective disinhibition of volitional motor plans facilitating improved movement vigor (*19*, *31*). Additionally, mDBS was associated with a progressive increase in resting-state M1-S1 beta coherence compared to cDBS. Prior studies have demonstrated that increased functional connectivity within the sensorimotor network is linked to improved motor control by enhancing the real-time integration of motor commands with somatosensory feedback (*47–49*). Enhanced sensorimotor cortical connectivity is also correlated with significant motor function improvements during stroke rehabilitation (*50–52*). Together, these findings suggest that mDBS may reinforce functional motor circuits involved in volitional movement, potentially explaining the observed cumulative antibradykinetic effect.

Our findings align with previous reports of plasticity-like effects in patients with PD. Specifically, Spencer et al. reported that a brief (∼40-s), continuous train of bursts of pallidal stimulation could increase hand movement amplitudes during a motor task, with effects lasting for several minutes beyond the cessation of stimulation (*22*). This stimulation pattern also potentiated underactive inhibitory projections from the striatum, suggesting the induction of LTP. However, similar bursts of subthalamic stimulation failed to elicit synaptic plasticity in that study. Conversely, Cavallo et al. demonstrated that subthalamic stimulation can induce plasticity-like effects, provided it is delivered in a highly specific, state-dependent manner rather than as a continuous train (*24*). In their work, administering STN DBS exclusively during fast wrist movements in a tablet-based motor task led to improved movement speeds in post-stimulation trials (lasting a few minutes).

Our study expands upon these previous findings by suggesting a more sustained therapeutic impact during daily activities beyond brief, structured experimental conditions. In the chronic validation phase, the progressive improvement in bradykinesia over hours under mDBS and its gradual reversal upon switching to cDBS, suggests the involvement of neural plasticity mechanisms within the cortico-basal motor network (*53*). The progressive reduction in symptoms over several hours is potentially consistent with late long-term potentiation (L-LTP), a robust form of activity-dependent neural plasticity (*54–56*). Additionally, the significant worsening of bradykinesia severity during the first two hours following the switch from mDBS to cDBS suggests that movement-responsive stimulation may strengthen neural circuits involved in volitional motion. However, subsequent indiscriminate, constant-amplitude stimulation may weaken these newly reinforced motor pathways, potentially reversing the induced plasticity (*23*, *24*).

Additionally, we developed a fully embedded classifier that utilized cortical biomarkers, such as alpha-beta MRD and broadband gamma MRS, to decode naturalistic forearm movement without external kinematic sensors. These neural signatures correlate with motor effort and extend to movements of other effectors, such as lower limbs during gait and the trunk during postural adjustments. Consequently, our neural decoder likely possessed a more generalizable capability for decoding contralateral movement and motor effort which may have contributed to the cumulative antibradykinetic effect observed with mDBS in the naturalistic environment.

### Limitations and future directions

Several caveats, however, warrant consideration. First, our evidence for neural plasticity is based on changes in symptom severity rather than a direct investigation of synaptic changes. Confirming the specific plasticity mechanisms involved will require direct physiological measurements, such as changes in evoked potentials, imaging studies or molecular markers of synaptic strengthening (*9*, *22*, *54*, *55*). Second, while subjects were monitored for up to 6 months under mDBS, the study was not powered to assess longer-term improvements or evaluate the persistence of clinical improvements after the cessation of stimulation. Despite these limitations, our findings suggest that mDBS may be able to enhance the therapeutic efficacy of DBS beyond acute symptomatic relief by providing cumulative motor improvements over time. These findings have implications for treatment strategies, potentially shifting the focus from merely titrating stimulation amplitude to manage symptoms and side effects toward refining stimulation patterns to strengthen motor pathway efficacy. This approach aligns with principles of motor learning and neurorehabilitation, wherein activity-dependent plasticity plays a central role in recovery (*24*, *57*).

Cortical signals have previously demonstrated higher predictive power for movement decoding than subcortical signals, even across varying stimulation amplitudes (*21*, *32*). This is consistent with the role of the sensorimotor cortex in motor planning and execution, which is upstream of basal ganglia physiology (*58*). It also aligns with prior work linking the excessive synchronization in the subthalamic beta band, associated with PD-related bradykinesia, to impaired movement decoding performance in STN (*59*). However, cortical sensing remains investigational, and the implantation of subdural electrodes, as used in this study, increases the invasiveness of the surgery (*16*, *27*). Nevertheless, these results collectively support the integration of cortical sensing with next-generation chronic, implantable devices, potentially using less invasive electrodes (*60*).

Our deployment period of 3–6 months is still likely insufficient for fully evaluating the long-term stability and benefits of mDBS. Considering that PD progresses over several years to decades, algorithms must be engineered to maintain robustness over these extended timescales. Future research must address challenges such as the evolving nature of neural biomarkers due to disease progression, changes in medication regimens, and lifestyle factors that may influence behavioral patterns (*18*, *61*). Developing online learning algorithms that can adapt to gradual changes while maintaining stability amidst rapid fluctuations represents an important engineering challenge for ensuring the long-term efficacy of these systems (*62*).

The potential applicability of mDBS may extend to other movement disorders, such as stroke neurorehabilitation, where cDBS has already shown promise. Baker et al. found that in individuals with persistent, moderate-to-severe upper-extremity impairment after an ischemic stroke, stimulating the cerebellar dentate nucleus - when explicitly paired with structured physical rehabilitation - promotes functional reorganization of the ipsilesional cortex (*63*). This pairing also resulted in clinically meaningful improvements in distal motor function and arm ability. Consequently, combining movement-responsive stimulation with physical therapy could offer additional treatment benefits. By modulating stimulation specifically during the intensive, repetitive use of impaired motor functions, this approach may "prime" functional networks to be more susceptible to activity-dependent synaptic changes during physical rehabilitation (*64*, *65*).

The mDBS paradigm represents a promising evolution beyond conventional, constant-amplitude DBS for PD. By dynamically adjusting stimulation amplitude based on real-time decoding of movement-related cortical signals, this approach highlights the antibradykinetic effects of mDBS in both constrained and naturalistic settings. Specifically, during structured motor tasks, mDBS demonstrated improved forearm speeds and counteracted the motor slowing observed in the cDBS condition over the course of the task, suggesting a potential neural reinforcement effect. This motor improvement was accompanied by a cumulative increase in sensorimotor cortical movement-related beta desynchronization, resting-state beta power and beta coherence during the motor tasks, identifying these as potential electrophysiological biomarkers of neural reinforcement. Further, during unconstrained daily activities, mDBS mitigated bradykinesia severity without worsening hyperkinetic side effects. The cumulative and reversible nature of the therapeutic benefit observed over hours suggests that mDBS may offer more than symptom masking; it could potentially provide sustained motor improvements by selectively reinforcing functional motor circuits. Consequently, this study positions mDBS as a strategy that integrates closed-loop neuromodulation and precise neural decoding of volitional motion, offering a more nuanced and potentially restorative treatment option for disorders of movement.

## Materials and Methods

### Patient enrollment and device implantation

Four male patients (mean age ± SEM: 60 ± 4 years) with idiopathic Parkinson’s disease (PD) were enrolled from a previous study cohort (ClinicalTrials.gov: NCT03582891) after providing informed consent, with protocols approved by the University of California, San Francisco Institutional Review Board. The inclusion criteria required persistent PD-related motor symptoms, prior responsiveness to levodopa, and candidacy for subthalamic deep brain stimulation (STN DBS). Each patient underwent bilateral implantation of the Medtronic^TM^ Summit RC+S investigational device as part of a prior clinical trial (*16*, *26*). Specifically, quadripolar DBS leads (Medtronic^TM^ 3389) were stereotactically placed in STN as per normal clinical implantation. Additionally, four-contact electrocorticography (ECoG) paddle leads (Medtronic^TM^ model 0913025) were positioned subdurally over the central sulcus, ensuring at least one contact covered the precentral gyrus and one covered the postcentral gyrus. The leads were connected to an investigational sensing-enabled implantable pulse generator (IPG; Medtronic^TM^ Summit RC+S model B35300R) placed in an infraclavicular pocket over the pectoralis muscle bilaterally. Each IPG was connected only to the ipsilateral leads. This device enabled chronic bidirectional communication: sensing local field potentials (LFPs) from STN and ECoG contacts while delivering programmable stimulation through STN contacts. Further surgical details are available in a previous publication by our group (*27*).

### Overview of study pipeline to develop and evaluate mDBS paradigm

The study protocol consisted of four phases conducted entirely remotely in the patient’s own home and supported via video-conferencing. Prior to the study, patients were clinically optimized as part of routine clinical care by a movement disorder neurologist. In Phase 1 of the study, preliminary stimulation amplitudes for both movement-responsive DBS (mDBS) and conventional, constant-amplitude DBS (cDBS) policies were titrated and refined. Patients completed brief, structured motor tasks: a 2-minute screen-tapping game on a tablet computer that we developed using the lab.js toolbox. The game was divided into five consecutive sets, each consisting of a ∼10-second stationary period (where patients kept their hands still on their lap without any movement) followed by a ∼10-second mobile period (where patients tapped a randomly-appearing corner black square as quickly as possible). Neural and wrist accelerometry signals were recorded during these tasks. These data were used to develop a preliminary cortical movement decoder and mDBS algorithm, which was then embedded on the Summit RC+S system.

Phase 2 (blinded, *acute* mDBS validation) required patients to perform the motor tasks on the tablet computer under mDBS and two control conditions: cDBS, and inverse mDBS (where stimulation amplitude was reduced during detected movement rather than increased). Inverse mDBS was included to confirm that any beneficial effects of mDBS on forearm speed were due to movement state-dependent stimulation and not simply due to the intermittency of the stimulation. Data from both Phases 1 and 2 were then combined to retrain the mDBS algorithm, ensuring the training data included signals collected during changes in stimulation amplitude. Finally, the three policies were administered again in a blinded, randomized and counter-balanced sequence while patients performed the same motor tasks. This allowed for the precise assessment and comparison of mDBS algorithm performance, forearm speeds, and error rates across the three DBS conditions. Patients consumed their medication prior to each testing session, ensuring they were in the medication-ON state and that there was no medication-related confound.

Phase 3 (blinded, *chronic* mDBS validation) compared the effects of mDBS and cDBS, excluding the inverse mDBS policy due to the significant worsening of forearm speed observed in Phase 2. This reduced the required patient time commitment and improved statistical power to examine for our main effect of mDBS versus cDBS. During an initial refinement period, neural and accelerometry data were collected (∼6-hours) during unsupervised daily activities and used to re-train the mDBS algorithm to now decode naturalistic movement states. Patients then entered an 18-day blinded evaluation period (36 half-day sessions) where they alternately received each stimulation policy (mDBS or cDBS) for ∼6-hours. The clinical effects of each policy were compared using a wearable symptom severity monitor (PKG) and self-reported scores for symptoms and side-effects not captured by the PKG watches. Finally, Phase 4 involved continuous, open-label deployment (24-hr) of mDBS for 3–6 months in one (Pt-3 and Pt-4) or both (Pt-1 and Pt-2) hemispheres to assess for sustained clinical effects and the stability of the mDBS algorithm’s performance.

### Titration of stimulation amplitudes for mDBS and cDBS

Following prior optimization during routine clinical care, at the start of the study, stimulation amplitudes for both the mDBS and cDBS policies were titrated remotely by a movement disorder neurologist via video-conference (Phase 1). Adjustments to the cDBS amplitudes for this clinical study were minimal (mean change in cDBS amplitude ± SEM: 0.09 ± 0.07 mA). For the mDBS policy, optimization involved defining lower and upper amplitude limits. The lower limit was set in the OFF dopaminergic state to determine a minimum stimulation level that suppressed breakthrough hypodopaminergic symptoms, including tremor. Conversely, the upper limit was established in the ON dopaminergic state, to maximally manage hypokinetic symptoms while also preventing adverse side effects related to overstimulation, including dyskinesia and dysarthria. These stimulation levels were refined based on clinical examination, patient feedback and PKG wearable outputs while mDBS was administered in Phases 2, 3 and 4.

### Neural signal acquisition and preprocessing

Cortical and subcortical LFPs were sampled at 500 Hz. For STN recordings, LFPs were acquired in a bipolar configuration using contacts adjacent to the monopolar stimulating cathode, leveraging common-mode rejection to eliminate stimulation artifacts (*16*). When dual monopolar stimulation was selected during clinical programming (four out of eight hemispheres), the remaining two contacts were used for sensing. Cortical LFPs were recorded using nonoverlapping bipolar electrode pairs. Specifically, recordings from the anterior montage targeted the primary motor cortex (M1), while the posterior montage focused on the somatosensory cortex (S1).

The collected neural data were transmitted from the Summit RC+S device to a nearby telemetry module and subsequently to a Microsoft Windows Surface Go tablet before being encrypted and uploaded to a secure cloud environment. The tablet was equipped with custom software that was built on the Summit RC+S application and in compliance with FDA regulations (CFR 820.30). The custom software is publicly available through the OpenMind Consortium (https://github.com/openmind-consortium). Initial preprocessing of the recorded neural signals was performed in MATLAB R2022b using the Analysis-rcs-data toolbox (*66*). Power spectral densities (PSDs) were calculated using Welch’s method, with ∼0.5-second epochs (256 samples), Hanning windows, and a 20% overlap (100-ms steps). This approach provided a frequency resolution of 2 Hz within the 0–100 Hz range.

### Movement labeling using wrist accelerometry measurements

Patients wore Apple Watch Series SE (2nd generation) devices on both wrists to record triaxial accelerometry data at a sampling frequency of 50 Hz. The Summit RC+S IPG also had an internal accelerometer with a sampling frequency of 64 Hz. To synchronize each Watch–IPG data pair, patients tapped their chest over the IPG with their contralateral hand, creating a time-locked artifact in both accelerometry signals that was used for timestamp realignment (*20*). To align neural recordings with the tablet-based screen-tapping task, Network Time Protocol (NTP) computer clock timestamps were synchronized across both data streams (*67*). During the acute validation phase, the patients performed structured motor tasks that generated data for distinguishing between mobile and stationary states. The accelerometry data collected during these tasks exhibited a bimodal distribution. For each patient and hemisphere, a movement threshold was determined by calculating the average of the two peak acceleration values from these distributions. This threshold was then applied during the acute and chronic validation phases to classify movement states. Epochs were labeled as ‘mobile’ if the acceleration magnitude at the end of the epoch exceeded the threshold, and as ‘stationary’ otherwise (*21*).

### Feature selection algorithms

Movement-related biomarkers were identified using Cohen’s *d* effect size, which quantified the signed difference in PSDs between mobile and stationary states and was normalized by the pooled standard deviation to account for class imbalance (*28*). This analysis provided a robust measure of the magnitude and direction of spectral power changes associated with movement. Positive Cohen’s *d* values indicated movement-related synchronization, while negative values represented movement-related desynchronization.

The mDBS algorithm was designed to be embedded on the Summit RC+S system, which supports up to four non-overlapping power bands (PBs) as features for binary classification (*20*). Consequently, we employed logistic regression with L2 regularization, which mitigates overfitting by penalizing large model weights, and used the synthetic minority over-sampling technique (SMOTE) to reduce class imbalance in our training datasets (*68*). Cortical PB features from S1 and M1 were prioritized due to their higher movement-predictive power relative to subcortical signals (*21*, *32*). To select these cortical PBs, we implemented the Cohen’s *d* initialization (CDI) algorithm. CDI identified the largest contiguous frequency ranges showing positive and negative Cohen’s *d* values, corresponding to MRS and MRD, respectively. Using Bayesian optimization (25 iterations, Tree-structured Parzen Estimator sampler) with five-fold cross-validation, we then selected the four optimal cortical PB features, constraining each feature’s frequency range to fall within an identified MRS or MRD band.

We evaluated CDI against two baseline feature selection methods that did not constrain power band frequency ranges: sequential feature selection (SFS) and iterative sequential optimization (ISO) (*33*). SFS iteratively added individual power bands (alternating between S1 and M1) that optimized cross-validation balanced accuracy until reaching four features or a performance plateau. In contrast, ISO identified all four PBs simultaneously within each iteration of Bayesian optimization. Algorithm performance was evaluated based on the cross-validated balanced accuracy of the resulting logistic regression models and overall processing runtime.

### Temporal smoothing hyperparameter optimization

Logistic regression models were trained on the power bands selected by each algorithm, incorporating three temporal smoothing hyperparameters native to the Summit RC+S system to mitigate spurious state transitions from transient neural fluctuations: 1) Onset duration (0.1–0.3 s): Required time in the mobile state to trigger a stimulation increase, 2) Termination duration (0.1–0.3 s): Required time in the stationary state to trigger a stimulation decrease, 3) State-change blanking duration (0.1–0.5 s): Refractory period following any state transition. These hyperparameters were tuned via Bayesian optimization (25 iterations, Tree-structured Parzen Estimator sampler) to maximize cross-validation balanced accuracy. For subsequent clinical evaluation, we selected the combination of four power bands and smoothing hyperparameters with optimal model performance.

### Implementation of movement state classifier for closed-loop control

The mDBS closed-loop algorithm was embedded on the Summit RC+S device. Movement state predictions were updated every 100 ms using the preceding 500 ms of neural data. Under mDBS, the stimulation amplitude was increased from its lower to upper limit upon detecting a mobile state. This increase followed a programmable linear ramp over 500 ms, though this ramp was extended if required to reduce paresthesia. The same ramp rate was used when transitioning from the mobile to the stationary state. Stimulation frequency (130 Hz or 150 Hz) and pulse width (60 μs), determined during routine clinical care by a movement disorders neurologist, were held constant throughout the study.

### mDBS algorithm performance metrics

The mDBS algorithm’s performance was assessed using balanced accuracy, comparing its predicted movement states to ground-truth labels derived from wrist accelerometry measurements. To assess the responsiveness of the algorithm, we calculated latency, defined as the time delay between the actual and detected movement state changes. Specifically, we computed the average time by which the predicted movement states, determined via Apple Watches, either lagged (positive latency values) or preceded (negative latency values) ground truth transitions. To prevent noise-induced underestimation of latency, ground-truth state transitions were required to persist for at least 0.5 seconds to be labeled as true state changes (*20*). Although raw accelerometry was unavailable from the PKG watches, the Summit RC+S device recorded mDBS-detected movement states at 1 Hz. These data allowed us to calculate the proportion of time spent in each state under active mDBS. Given the algorithm’s rapid timescale and the onboard storage capacity of the RC+S device, each log file contained data spanning up to 12 hours. To detect potential performance variations caused by neural biomarker drift or recording artifacts, we evaluated changes in movement state-time proportions across the open-label deployment phase.

### Metrics for comparing mDBS with cDBS and inverse mDBS control policies

In the acute validation phase, the impact of mDBS on motor performance was evaluated by measuring inter-tap intervals (a measure of forearm motor speed) during the tablet-based, screen-tapping game. Error rates (taps outside a 20-pixel margin of each 50 x 50 target black square) during the task were measured to check for increased involuntary, dyskinetic movements from over-stimulation. In the chronic validation phase, patients wore bilateral PKG watches, which recorded triaxial accelerometry to derive 2-minute epoch scores for bradykinesia, dyskinesia, and tremor severity using validated algorithms (*37–39*). The PKG bradykinesia score served as the primary metric for comparison between mDBS and cDBS. Secondary outcomes included twice-daily self-administered questionnaires (REDCap) completed after each DBS policy administration. On these REDCap surveys, patients rated the severity of PD symptoms and side-effects not captured by the PKG watches (dystonia, gait, dysarthria) on a 0-10 visual analog scale that were then converted to a percentage scale. Finally, the open-label deployment phase involved the continuous implementation of the mDBS algorithm (in one or both hemispheres) during unconstrained daily life. Patients continued to complete weekly REDCap surveys assessing the severity of bradykinesia and dyskinesia, as well as their sleep quality.

### Statistical and computational analyses

To evaluate the outcome variables across groups while accounting for the interdependence between hemispheres within the same patient, we used linear mixed models (LMMs). In these models, the DBS policy was treated as a fixed effect, while patients and their corresponding hemispheres were treated as random effects. Our primary hypotheses focused on analyzing the effects of mDBS on forearm movement speeds and error rates during the acute validation phase, as well as bradykinesia and dyskinesia scores during the chronic validation phase. Since these group-level analyses (using LMMs) were designed to test predefined hypotheses, we did not apply false discovery rate (FDR) correction. However, p-values from the comparisons of secondary self-reported symptom scores in the chronic validation phase - dystonia, gait, and dysarthria - were subjected to FDR correction to adjust for multiple comparisons. Individual analysis of the therapeutic effects of mDBS compared to cDBS in the chronic validation phase was performed using independent *t*-tests. All statistical tests were two-tailed, with a significance threshold of α = 0.05.

Additionally, we calculated the Cohen’s *d* effect size for PSDs between 0 and 100 Hz for each brain region. To identify frequency ranges with significant Cohen’s *d* effect sizes, we performed one-sample *t*-tests (with a population mean of 0) followed by FDR correction for multiple frequency comparisons. LMMs were then used to compare the absolute, average Cohen’s *d* effect sizes across brain regions to assess their movement-predictive power. We also used LMMs to evaluate the performance of feature selection algorithms (CDI, SFS, and ISO) before and after hyperparameter tuning, as well as to compare the processing runtimes of these algorithms. Further, in the chronic validation phase, LMMs were employed to assess the performance of the mDBS algorithm (balanced accuracy and latency) before and after the 18-day evaluation period, to evaluate the time spent in each movement state under mDBS and to compare the average stimulation amplitudes administered under mDBS versus cDBS.

To evaluate potential neural plasticity effects induced by mDBS, we analyzed progressive changes in behavioral performance and neural activity. In the acute validation phase, we calculated median inter-tap intervals for each of the five mobile periods in the screen-tapping task. Pearson *r* correlation coefficients were computed across the five sets under mDBS, cDBS, and inverse mDBS conditions and compared using LMMs. Similar correlation analyses were performed on neural metrics across sets. Movement-related desynchronization was assessed by calculating Cohen’s *d* effect size for M1 beta power across the five sets, with a ±1-second buffer between sets (including both stationary and mobile periods). Resting-state (stationary periods) and mobile-state (movement periods) M1 beta power, as well as M1–S1 beta coherence, were extracted per set. Pearson *r* correlation coefficients across sets were evaluated and compared across DBS policies using LMMs. In the chronic validation, Pearson *r* correlation coefficients were calculated to assess cumulative changes in PKG bradykinesia scores over time (in minutes) under mDBS compared to cDBS. Data within a ±30-minute window before and after each scheduled DBS policy switch were excluded to account for potential variability in the switch time due to patient availability.

Analyses were performed using several Python packages. Classical statistical tests were performed using SciPy, machine learning models were developed with scikit-learn and LMMs were fitted with pymer4. Bayesian optimization was conducted using Optuna, and functional connectivity was quantified via mne-connectivity. High-resolution time-frequency spectrograms were generated using the superlet transform algorithm (*69*). Additionally, rcssim - an open-source package developed by our group - was employed to simulate the computational processes of the Summit RC+S device and tailor mDBS algorithm development to the device’s hardware specifications (https://github.com/Weill-Neurohub-OPTiMaL/rcs-simulation). Further information on the rcssim package can be found in a prior publication (*20*).

## Data Availability

All de-identified, processed data and relevant code used to generate figures and perform analyses described in this manuscript can be made available upon request, in compliance with institutional and regulatory requirements.

## Acknowledgments

We thank our patient cohort for their involvement in this study. We also thank Colin Hoy for his help in preparing Figure 1B. In addition, Figures 1D and 2B were designed using elements from Flaticon.com.

## Author contributions

Conceptualization: SJL, DJL, JAH

Supervision: SJL, PAS

Methodology: DJL, SJL, PAS

Investigation: DJL, JS, VC

Funding acquisition: SJL, JAH

Writing - original draft: DJL, SJL, PAS

Writing - review & editing: DJL, SJL, PAS, JAH, VC, JS

Project administration - DJL, SJL

## Computing interests

PAS receives funding from Medtronic, Inc. and Boston Scientific Inc. for salary support of clinical fellows. PAS is a consultant for INBRAIN Neuroelectronics Inc and Echo Neurotechnologies. SJL was previously a consultant for Iota Biosciences and is co-founder and CEO of Ocean Neuro. DJL is a part-time contractor for Echo Neurotechnologies. PAS, SJL, and DJL are listed inventors on the patent US2024/041516 which relates to daytime decoding of movement for adaptive deep brain stimulation in neurological disorders, including Parkinson’s disease and stroke.

## Supplementary Materials

**Fig. S1.**
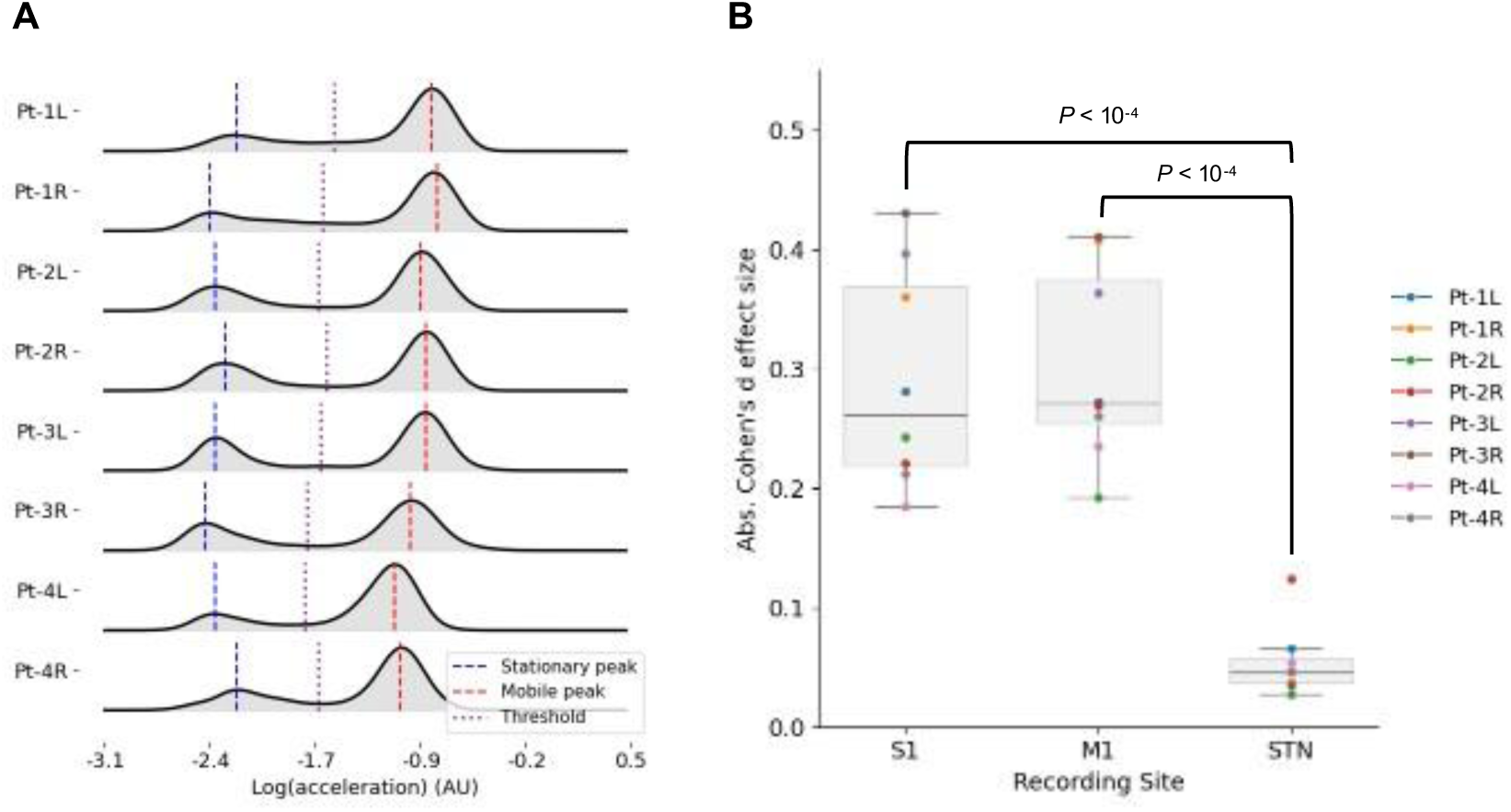
(A) Distributions of accelerometry measurements from all patients and bilateral forearms. Stationary and mobile peaks from each bimodal distribution were labeled. The average acceleration value from the two peaks in each distribution was computed and set as the threshold to delineate between binary movement states. (B) Absolute, average Cohen’s *d* effect size from each hemisphere and brain region was computed within 0 – 100 Hz.

**Fig. S2.**
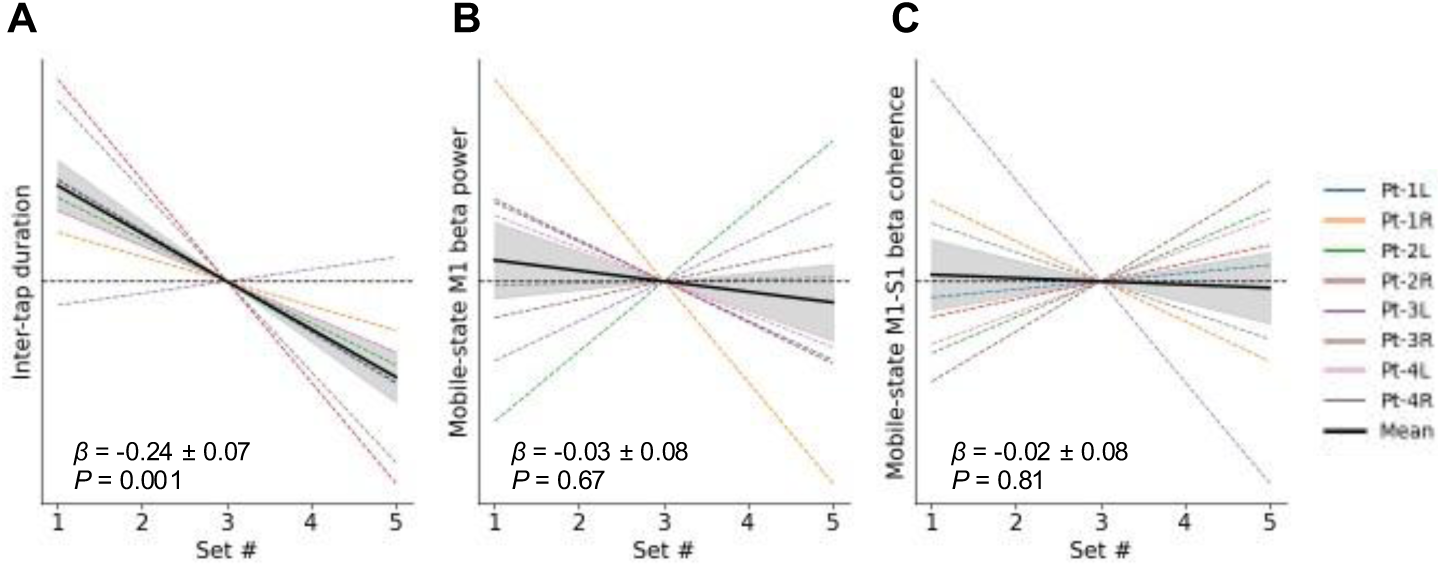
Pearson *r* correlation coefficients modeled the progressive variation in (A) inter-tap duration, (B) median mobile-state M1 beta power (during only mobile periods), and (C) M1-S1 mobile-state beta coherence over the five sets of the screen-tapping game under mDBS compared to cDBS. In (A - C), the *r* correlation coefficients for cDBS were subtracted from those for mDBS for normalization. The median *r* correlation coefficient (after normalization) for each patient and hemisphere was plotted as a regression line. The group-level average regression line (solid black) and the standard error of the mean (grey shading) were also included. Statistical comparisons between mDBS and cDBS were performed on the *r* correlation coefficients (without normalization) using LMMs. Patients and hemispheres were included as random effects in each LMM. L and R indicate left and right forearms used by patients. L and R indicate left and right forearms used by patients.

**Fig. S3.**
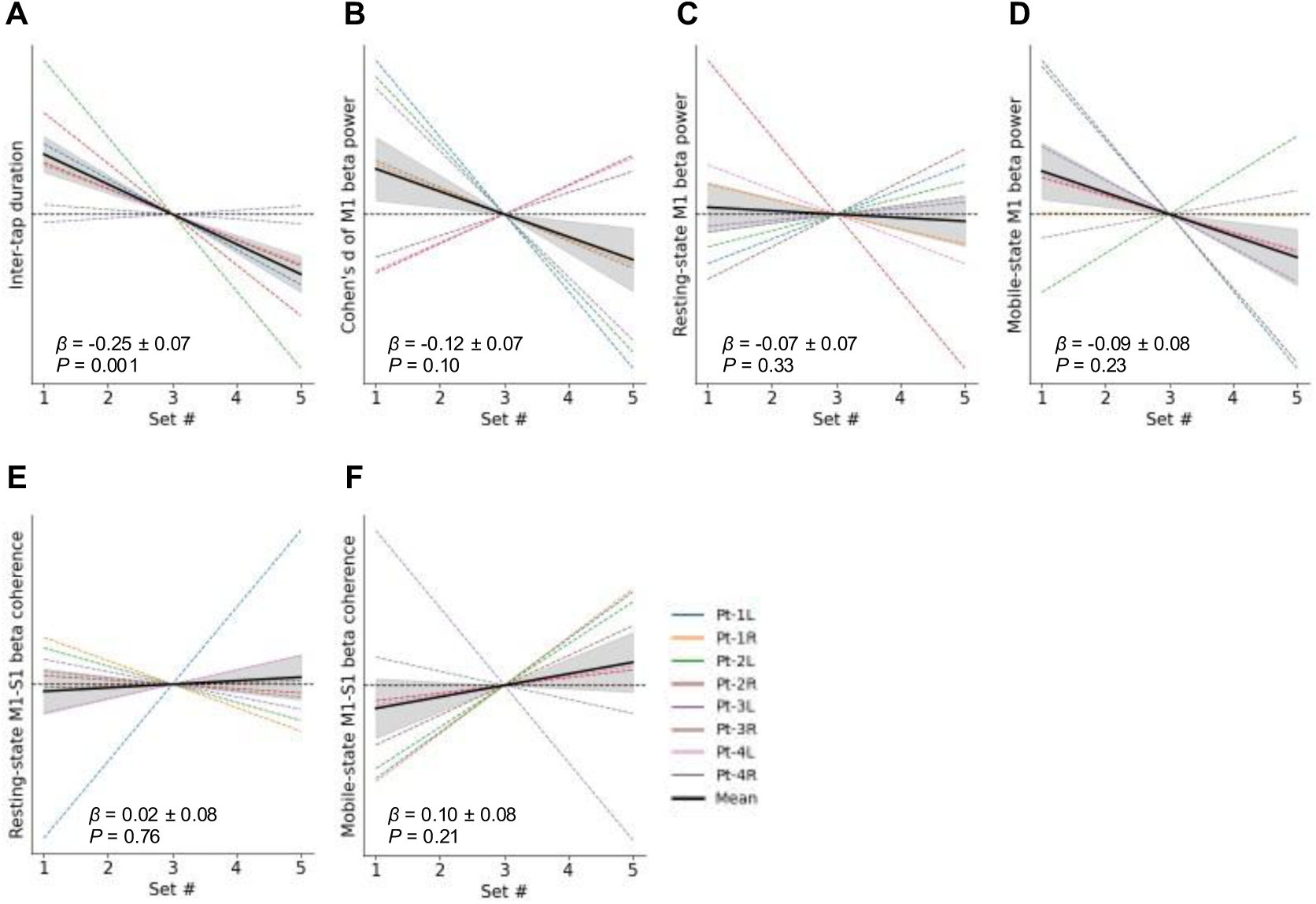
Pearson *r* correlation coefficients modeled the progressive variation in (A) inter-tap duration, (B) Cohen’s *d* effect size of M1 beta power (used to quantify movement-related desynchronization), (C) median resting-state M1 beta power (during only stationary periods), (D) median mobile- state M1 beta power (during only mobile periods), (E) M1-S1 resting-state beta coherence and (F) M1-S1 mobile-state beta coherence over the five sets of the screen-tapping game under mDBS compared to cDBS. In (A - F), the *r* correlation coefficients for cDBS were subtracted from those for mDBS for normalization. The median *r* correlation coefficient (after normalization) for each patient and hemisphere was plotted as a regression line. The group-level average regression line (solid black) and the standard error of the mean (grey shading) were also included. Statistical comparisons between mDBS and cDBS were performed on the *r* correlation coefficients (without normalization) using LMMs. Patients and hemispheres were included as random effects in each LMM. L and R indicate left and right forearms used by patients. L and R indicate left and right forearms used by patients.

**Fig. S4.**
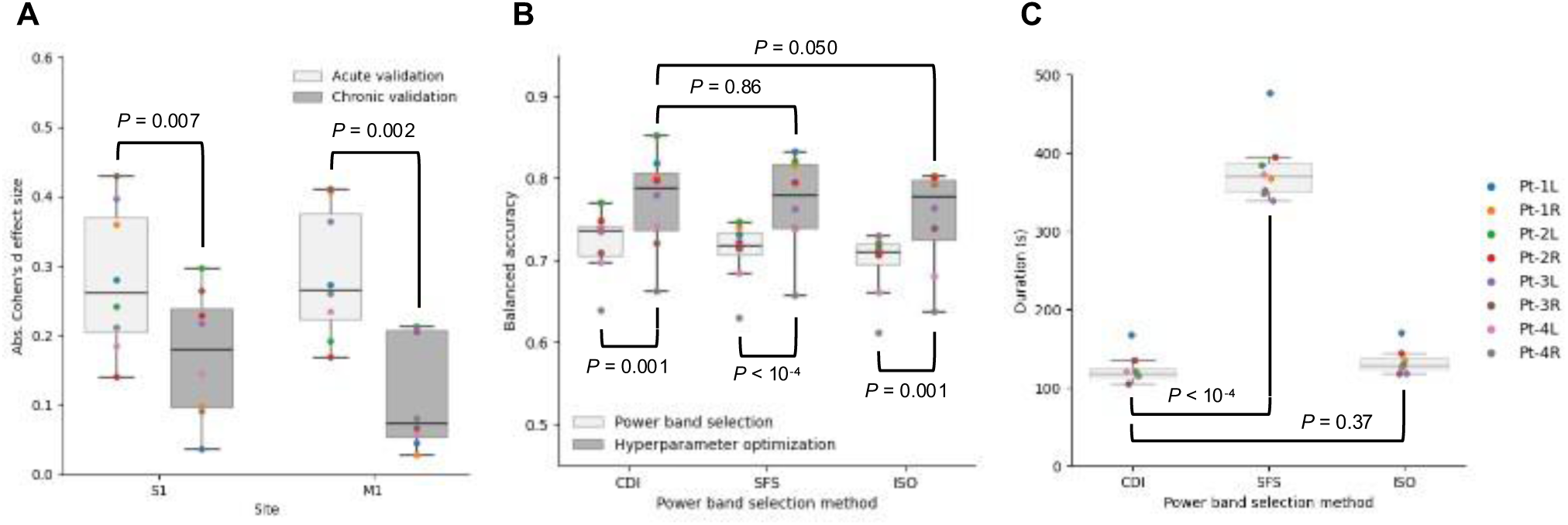
(A) For each patient and hemisphere, the average, absolute Cohen’s *d* effect size (for S1 and M1) was computed within 0 – 100 Hz. (B) Comparison of PB feature selection algorithms (CDI, SFS and ISO) using linear mixed models (LMMs). Mean balanced accuracy from five-fold cross validation (CV) was computed for each hemisphere and used as the fixed effect in the LMMs. Bayesian optimization was applied to tune smoothing hyperparameters for modeling forearm movement dynamics. PBs yielding the highest CV performance were selected when the mDBS algorithm was re-trained for the chronic validation phase. (C) The processing times for each PB feature selection algorithm were compared using LMMs. For all LMMs used in this figure, patients and hemispheres were treated as random effects. L and R denote data from the left and right forearms of patients.

**Fig. S5.**
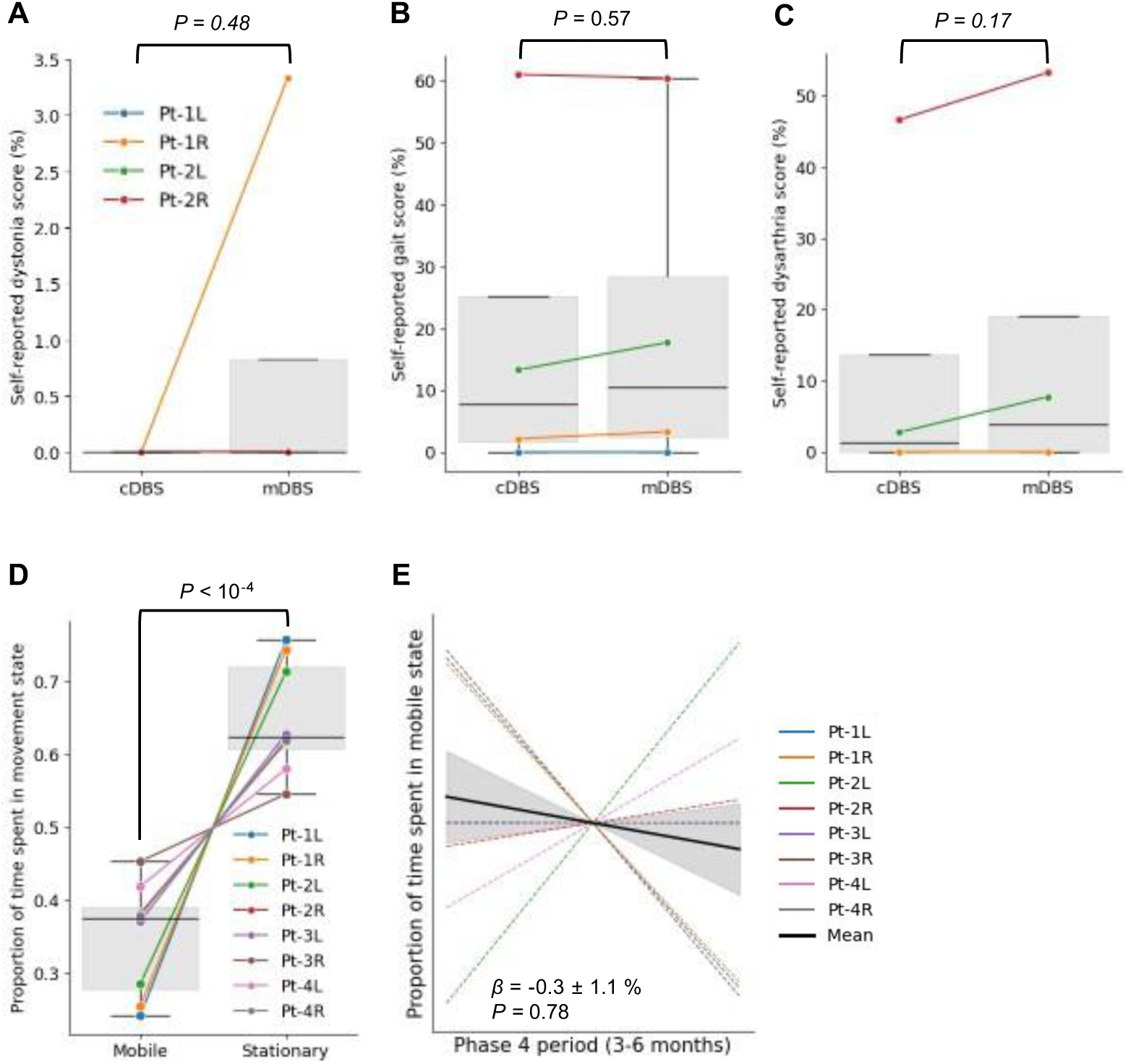
Linear mixed models (LMMs) were used to compare several secondary outcome measures during mDBS and cDBS administration: self-reported (A) dystonia, (B) gait and (C) dysarthria severity scores. The p-values from these LMMs underwent FDR correction for multiple comparisons. (D) The average proportion of time spent in each movement state (stationary vs mobile) under mDBS during the open-label deployment phase was computed from stimulation log files and quantified using LMMs. (E) Pearson *r* correlation coefficients were used to model the variation in proportion of time spent in the mobile state over the open-label deployment phase. In (D-E), the proportions of time spent in each movement state were analyzed only during awake hours. For each patient and hemisphere, the average regression line was plotted, along with the group-level average regression line (solid black). The standard error of the mean was shown as grey shading for the average regression line. For all LMMs used in this figure, patients and hemispheres were treated as random effects. L and R indicate left and right forearms used by patients.

**Table S1.** Deep brain stimulation parameters for each patient and hemisphere during constant-amplitude DBS and movement-responsive DBS.

| Patient ID | Hemisphere | Frequency (Hz) | Contacts | Pulse width (μs) |
| --- | --- | --- | --- | --- |
| Pt-1 | L | 130 | 2-C+ | 60 |
|  | R | 130 | 1-C+ | 60 |
| Pt-2 | L | 130 | 1-C+ | 60 |
|  | R | 130 | 1-3-C+ | 60 |
| Pt-3 | L | 130 | 2-3-C+ | 60 |
|  | R | 130 | 2-3-C+ | 60 |
| Pt-4 | L | 150 | 1-C+ | 60 |
|  | R | 150 | 1-2-C+ | 60 |

## References and Notes

1. N. G. Laxpati, W. S. Kasoff, R. E. Gross, Deep brain stimulation for the treatment of epilepsy: circuits, targets, and trials. Neurotherapeutics 11, 508–526 (2014).

2. P. E. Holtzheimer, H. S. Mayberg, Deep brain stimulation for psychiatric disorders. Annu. Rev. Neurosci. 34, 289–307 (2011).

3. G. Deuschl, C. Schade-Brittinger, P. Krack, J. Volkmann, H. Schäfer, K. Bötzel, C. Daniels, A. Deutschländer, U. Dillmann, W. Eisner, D. Gruber, W. Hamel, J. Herzog, R. Hilker, S. Klebe, M. Kloss, J. Koy, M. Krause, A. Kupsch, D. Lorenz, S. Lorenzl, H. M. Mehdorn, J. R. Moringlane, W. Oertel, M. O. Pinsker, H. Reichmann, A. Reuss, G.-H. Schneider, A. Schnitzler, U. Steude, V. Sturm, L. Timmermann, V. Tronnier, T. Trottenberg, L. Wojtecki, E. Wolf, W. Poewe, J. Voges, German Parkinson Study Group, Neurostimulation Section, A randomized trial of deep-brain stimulation for Parkinson’s disease. N. Engl. J. Med. 355, 896–908 (2006).

4. A. Kupsch, R. Benecke, J. Müller, T. Trottenberg, G.-H. Schneider, W. Poewe, W. Eisner, A. Wolters, J.-U. Müller, G. Deuschl, M. O. Pinsker, I. M. Skogseid, G. K. Roeste, J. Vollmer-Haase, A. Brentrup, M. Krause, V. Tronnier, A. Schnitzler, J. Voges, G. Nikkhah, J. Vesper, M. Naumann, J. Volkmann, Deep-Brain Stimulation for Dystonia Study Group, Pallidal deep-brain stimulation in primary generalized or segmental dystonia. N. Engl. J. Med. 355, 1978–1990 (2006).

5. G. Deuschl, J. Raethjen, H. Hellriegel, R. Elble, Treatment of patients with essential tremor. Lancet Neurol. 10, 148–161 (2011).

6. J. Herron, V. Kremen, J. D. Simeral, H. Dawes, G. A. Worrell, P. A. Starr, T. Denison, D. Borton, The convergence of neuromodulation and brain–computer interfaces. Nat. Rev. Bioeng. 2, 628–630 (2024).

7. S. E. Cooper, A. M. Noecker, H. Abboud, J. L. Vitek, C. C. McIntyre, Return of bradykinesia after subthalamic stimulation ceases: relationship to electrode location. Exp. Neurol. 231, 207–213 (2011).

8. Z. Keresztenyi, P. Valkovic, T. Eggert, U. Steude, J. Hermsdörfer, J. Laczko, K. Bötzel, The time course of the return of upper limb bradykinesia after cessation of subthalamic stimulation in Parkinson’s disease. Parkinsonism Relat. Disord. 13, 438–442 (2007).

9. T. A. Spix, S. Nanivadekar, N. Toong, I. M. Kaplow, B. R. Isett, Y. Goksen, A. R. Pfenning, A. H. Gittis, Population-specific neuromodulation prolongs therapeutic benefits of deep brain stimulation. [Preprint] (2025).

10. P. Temperli, J. Ghika, J.-G. Villemure, P. R. Burkhard, J. Bogousslavsky, F. J. G. Vingerhoets, How do parkinsonian signs return after discontinuation of subthalamic DBS? [Preprint] (2026). 10.1212/wnl.60.1.78.

11. H. Teymourian, F. Tehrani, K. Longardner, K. Mahato, T. Podhajny, J.-M. Moon, Y. G. Kotagiri, J. R. Sempionatto, I. Litvan, J. Wang, Closing the loop for patients with Parkinson disease: where are we? Nat. Rev. Neurol. 18, 497–507 (2022).

12. H. Cagnan, T. Denison, C. McIntyre, P. Brown, Emerging technologies for improved deep brain stimulation. Nat. Biotechnol. 37, 1024–1033 (2019).

13. C. E. Bouton, A. Shaikhouni, N. V. Annetta, M. A. Bockbrader, D. A. Friedenberg, D. M. Nielson, G. Sharma, P. B. Sederberg, B. C. Glenn, W. J. Mysiw, A. G. Morgan, M. Deogaonkar, A. R. Rezai, Restoring cortical control of functional movement in a human with quadriplegia. Nature 533, 247–250 (2016).

14. M. S. Willsey, N. P. Shah, D. T. Avansino, N. V. Hahn, R. M. Jamiolkowski, F. B. Kamdar, L. R. Hochberg, F. R. Willett, J. M. Henderson, A high-performance brain-computer interface for finger decoding and quadcopter game control in an individual with paralysis. Nat. Med. 31, 96–104 (2025).

15. E. Opri, S. Cernera, R. Molina, R. S. Eisinger, J. N. Cagle, L. Almeida, T. Denison, M. S. Okun, K. D. Foote, A. Gunduz, Chronic embedded cortico-thalamic closed-loop deep brain stimulation for the treatment of essential tremor. Sci. Transl. Med. 12, eaay7680 (2020).

16. C. R. Oehrn, S. Cernera, L. H. Hammer, M. Shcherbakova, J. Yao, A. Hahn, S. Wang, J. L. Ostrem, S. Little, P. A. Starr, Chronic adaptive deep brain stimulation versus conventional stimulation in Parkinson’s disease: a blinded randomized feasibility trial. Nat. Med. 30, 3345–3356 (2024).

17. S. Little, A. Pogosyan, S. Neal, B. Zavala, L. Zrinzo, M. Hariz, T. Foltynie, P. Limousin, K. Ashkan, J. FitzGerald, A. L. Green, T. Z. Aziz, P. Brown, Adaptive deep brain stimulation in advanced Parkinson disease: Adaptive DBS in PD. Ann. Neurol. 74, 449–457 (2013).

18. H. M. Bronte-Stewart, M. Beudel, J. L. Ostrem, S. Little, L. Almeida, A. Ramirez-Zamora, A. Fasano, T. Hassell, K. T. Mitchell, E. Moro, M. Gostkowski, G. Chattree, R. M. A. de Bie, M. de Neeling, D. Piña-Fuentes, B. Swinnen, P. A. Starr, L. H. Hammer, K. D. Foote, R. M. Richardson, A. Flaherty, A. Boogers, Q. Sa’di, S. Meoni, A. Castrioto, S. Stanslaski, R. L. S. Summers, L. Tonder, Y. Tan, H. Berrier, T. J. Goble, R. S. Raike, T. M. Herrington, ADAPT-PD Investigators, Long-term personalized adaptive deep brain stimulation in Parkinson disease: A nonrandomized clinical trial: A nonrandomized clinical trial. JAMA Neurol. 82, 1171–1180 (2025).

19. A. A. Kühn, F. Kempf, C. Brücke, L. Gaynor Doyle, I. Martinez-Torres, A. Pogosyan, T. Trottenberg, A. Kupsch, G.-H. Schneider, M. I. Hariz, W. Vandenberghe, B. Nuttin, P. Brown, High-frequency stimulation of the subthalamic nucleus suppresses oscillatory beta activity in patients with Parkinson’s disease in parallel with improvement in motor performance. J. Neurosci. 28, 6165–6173 (2008).

20. T. C. Dixon, G. Strandquist, A. Zeng, T. Frączek, R. Bechtold, D. Lawrence, S. Ravi, P. A. Starr, J. L. Gallant, J. A. Herron, S. J. Little, Movement-responsive deep brain stimulation for Parkinson’s disease using a remotely optimized neural decoder. *Nat*. Biomed. Eng., doi: 10.1038/s41551-025-01438-0 (2025).

21. D. Lawrence, G. Avraham, J. Yao, L. Li, C. Shi, P. A. Starr, S. J. Little, Cortico-basal oscillations index naturalistic movements during deep brain stimulation. Brain, doi: 10.1093/brain/awaf466 (2025).

22. K. A. Spencer, A. Boogers, S. Sumarac, D. B. J. Crompton, L. A. Steiner, L. Zivkovic, Y. Buren, A. Boutet, A. M. Lozano, S. K. Kalia, W. D. Hutchison, A. Fasano, L. Milosevic, Modulating inhibitory synaptic plasticity to restore basal ganglia dynamics in Parkinson’s disease. Brain 148, 2299–2305 (2025).

23. E. A. Yttri, J. T. Dudman, Opponent and bidirectional control of movement velocity in the basal ganglia. Nature 533, 402–406 (2016).

24. A. Cavallo, R. M. Köhler, J. L. Busch, J. G. V. Habets, T. Merk, P. Zvarova, J. Vanhoecke, T. S. Binns, B. Al-Fatly, A. L. de Almeida Marcelino, N. Darcy, G.-H. Schneider, P. Krause, A. Horn, K. Faust, D. M. Herz, E. Yttri, H. Cagnan, A. A. Kühn, W.-J. Neumann, Differential modulation of movement speed with state-dependent deep brain stimulation in Parkinson’s disease. Sci. Adv. 11, eadx6849 (2025).

25. J. E. Markowitz, W. F. Gillis, M. Jay, J. Wood, R. W. Harris, R. Cieszkowski, R. Scott, D. Brann, D. Koveal, T. Kula, C. Weinreb, M. A. M. Osman, S. R. Pinto, N. Uchida, S. W. Linderman, B. L. Sabatini, S. R. Datta, Spontaneous behaviour is structured by reinforcement without explicit reward. Nature 614, 108–117 (2023).

26. R. ’ee Gilron, S. Little, R. Perrone, R. Wilt, C. de Hemptinne, M. S. Yaroshinsky, C. A. Racine, S. S. Wang, J. L. Ostrem, P. S. Larson, D. D. Wang, N. B. Galifianakis, I. O. Bledsoe, M. San Luciano, H. E. Dawes, G. A. Worrell, V. Kremen, D. A. Borton, T. Denison, P. A. Starr, Long-term wireless streaming of neural recordings for circuit discovery and adaptive stimulation in individuals with Parkinson’s disease. Nat. Biotechnol. 39, 1078–1085 (2021).

27. N. C. Swann, C. de Hemptinne, S. Miocinovic, S. Qasim, J. L. Ostrem, N. B. Galifianakis, M. S. Luciano, S. S. Wang, N. Ziman, R. Taylor, P. A. Starr, Chronic multisite brain recordings from a totally implantable bidirectional neural interface: experience in 5 patients with Parkinson’s disease. J. Neurosurg. 128, 605–616 (2018).

28. D. Lakens, Calculating and reporting effect sizes to facilitate cumulative science: a practical primer for t-tests and ANOVAs. Front. Psychol. 4, 863 (2013).

29. E. D. Kondylis, M. J. Randazzo, A. Alhourani, W. J. Lipski, T. A. Wozny, Y. Pandya, A. S. Ghuman, R. S. Turner, D. J. Crammond, R. M. Richardson, Movement-related dynamics of cortical oscillations in Parkinson’s disease and essential tremor. Brain 139, 2211–2223 (2016).

30. N. E. Crone, D. L. Miglioretti, B. Gordon, R. P. Lesser, Functional mapping of human sensorimotor cortex with electrocorticographic spectral analysis. II. Event-related synchronization in the gamma band. Brain 121 **( Pt** **12**), 2301–2315 (1998).

31. A. A. Kühn, D. Williams, A. Kupsch, P. Limousin, M. Hariz, G.-H. Schneider, K. Yarrow, P. Brown, Event-related beta desynchronization in human subthalamic nucleus correlates with motor performance. Brain 127, 735–746 (2004).

32. T. Merk, V. Peterson, W. J. Lipski, B. Blankertz, R. S. Turner, N. Li, A. Horn, R. M. Richardson, W.-J. Neumann, Electrocorticography is superior to subthalamic local field potentials for movement decoding in Parkinson’s disease. Elife 11 (2022).

33. D. W. Aha, R. L. Bankert, “A comparative evaluation of sequential feature selection algorithms” in Learning from Data (Springer New York, New York, NY, 1996)Lecture Notes in Statistics, pp. 199–206.

34. J. R. Manning, J. Jacobs, I. Fried, M. J. Kahana, Broadband shifts in local field potential power spectra are correlated with single-neuron spiking in humans. J. Neurosci. 29, 13613–13620 (2009).

35. S. Sakhavi, C. Guan, S. Yan, Learning temporal information for brain-computer interface using convolutional neural networks. IEEE Trans. Neural Netw. Learn. Syst. 29, 5619–5629 (2018).

36. S. M. Peterson, S. H. Singh, N. X. R. Wang, R. P. N. Rao, B. W. Brunton, Behavioral and neural variability of naturalistic arm movements. eNeuro 8, ENEURO.0007–21.2021 (2021).

37. A. Santiago, J. W. Langston, R. Gandhy, R. Dhall, S. Brillman, L. Rees, C. Barlow, Qualitative evaluation of the personal KinetiGraphTM Movement Recording System in a Parkinson’s clinic. J. Parkinsons. Dis. 9, 207–219 (2019).

38. R. Joshi, J. M. Bronstein, A. Keener, J. Alcazar, D. D. Yang, M. Joshi, N. Hermanowicz, PKG movement recording system use shows promise in routine clinical care of patients with Parkinson’s disease. Front. Neurol. 10, 1027 (2019).

39. P. Farzanehfar, H. Woodrow, M. Braybrook, S. McGregor, A. Evans, F. Nicklason, M. Horne, Objective measurement in routine care of people with Parkinson’s disease improves outcomes. NPJ Parkinsons Dis. 4, 10 (2018).

40. M. M. McGregor, A. B. Nelson, Circuit mechanisms of Parkinson’s disease. Neuron 101, 1042–1056 (2019).

41. J. Y. Hansen, S. Cauzzo, K. Singh, M. G. García-Gomar, J. M. Shine, M. Bianciardi, B. Misic, Integrating brainstem and cortical functional architectures. Nat. Neurosci. 27, 2500–2511 (2024).

42. A. Parent, L. N. Hazrati, Functional anatomy of the basal ganglia. II. The place of subthalamic nucleus and external pallidum in basal ganglia circuitry. Brain Res. Brain Res. Rev. 20, 128–154 (1995).

43. O. Darbin, N. Hatanaka, S. Takara, N. Kaneko, S. Chiken, D. Naritoku, A. Martino, A. Nambu, Subthalamic nucleus deep brain stimulation driven by primary motor cortex γ2 activity in parkinsonian monkeys. Sci. Rep. 12, 6493 (2022).

44. P. Mazzoni, A. Hristova, J. W. Krakauer, Why don’t we move faster? Parkinson’s disease, movement vigor, and implicit motivation. J. Neurosci. 27, 7105–7116 (2007).

45. L. M. McDonald, H. J. Griffin, A. Angeli, M. Torkamani, D. Georgiev, M. Jahanshahi, Motivational modulation of self-initiated and externally triggered movement speed induced by threat of shock: Experimental evidence for paradoxical kinesis in Parkinson’s disease. PLoS One 10, e0135149 (2015).

46. R. S. Turner, M. Desmurget, Basal ganglia contributions to motor control: a vigorous tutor. Curr. Opin. Neurobiol. 20, 704–716 (2010).

47. T. Umeda, T. Isa, Y. Nishimura, The somatosensory cortex receives information about motor output. Sci. Adv. 5, eaaw5388 (2019).

48. C. L. Witham, M. Wang, S. N. Baker, Corticomuscular coherence between motor cortex, somatosensory areas and forearm muscles in the monkey. Front. Syst. Neurosci. 4, 38 (2010).

49. K. A. Edholm, M. C. Vinding, C. Pfeiffer, A. Svenningsson, E. Fransén, M. Sundgren, H. Sjöström, N. Edvall, D. Lundqvist, J. Waldthaler, Cortical response to proprioceptive stimulation in primary orthostatic tremor - a magnetoencephalography study. Clin. Neurophysiol. Pract. 10, 159–166 (2025).

50. M. L. Ingemanson, J. R. Rowe, V. Chan, E. T. Wolbrecht, D. J. Reinkensmeyer, S. C. Cramer, Somatosensory system integrity explains differences in treatment response after stroke. Neurology 92, e1098–e1108 (2019).

51. S. Vahdat, M. Darainy, A. Thiel, D. J. Ostry, A single session of robot-controlled proprioceptive training modulates functional connectivity of sensory motor networks and improves reaching accuracy in chronic stroke. Neurorehabil. Neural Repair 33, 70–81 (2019).

52. H. R. McGregor, P. L. Gribble, Functional connectivity between somatosensory and motor brain areas predicts individual differences in motor learning by observing. J. Neurophysiol. 118, 1235–1243 (2017).

53. D. D. Wang, C. de Hemptinne, Harnessing neuronal plasticity for sustained symptom relief with DBS. Trends Neurosci. 48, 827–828 (2025).

54. H. Hagena, D. Manahan-Vaughan, Differentiation in the protein synthesis-dependency of persistent synaptic plasticity in mossy fiber and associational/commissural CA3 synapses in vivo. Front. Integr. Neurosci. 7, 10 (2013).

55. M. A. Sutton, E. M. Schuman, Dendritic protein synthesis, synaptic plasticity, and memory. Cell 127, 49–58 (2006).

56. C. Gkogkas, N. Sonenberg, M. Costa-Mattioli, Translational control mechanisms in long-lasting synaptic plasticity and memory. J. Biol. Chem. 285, 31913–31917 (2010).

57. K. Ganguly, M.-M. Poo, Activity-dependent neural plasticity from bench to bedside. Neuron 80, 729–741 (2013).

58. J. L. Lanciego, N. Luquin, J. A. Obeso, Functional neuroanatomy of the basal ganglia. Cold Spring Harb. Perspect. Med. 2, a009621 (2012).

59. S. Khawaldeh, G. Tinkhauser, S. A. Shah, K. Peterman, I. Debove, T. A. K. Nguyen, A. Nowacki, M. L. Lachenmayer, M. Schuepbach, C. Pollo, P. Krack, M. Woolrich, P. Brown, Subthalamic nucleus activity dynamics and limb movement prediction in Parkinson’s disease. Brain 143, 582–596 (2020).

60. J. D. Olson, J. D. Wander, L. Johnson, D. Sarma, K. Weaver, E. J. Novotny, J. G. Ojemann, F. Darvas, Comparison of subdural and subgaleal recordings of cortical high-gamma activity in humans. Clin. Neurophysiol. 127, 277–284 (2016).

61. S. Cernera, C. R. Oehrn, L. H. Hammer, M. Shcherbakova, J. Yao, A. Hahn, S. Wang, J. L. Ostrem, S. Little, P. A. Starr, Sustained clinical benefit of adaptive deep brain stimulation in Parkinson’s disease using gamma oscillations: A case report. Mov. Disord. 40, 345–350 (2025).

62. G. H. Wilson, E. A. Stein, F. Kamdar, D. T. Avansino, T. K. Pun, R. Gross, T. Hosman, T. Singer-Clark, A. Kapitonava, L. R. Hochberg, J. D. Simeral, K. V. Shenoy, S. Druckmann, J. M. Henderson, F. R. Willett, Long-term unsupervised recalibration of cursor-based intracortical brain-computer interfaces using a hidden Markov model. *Nat*. Biomed. Eng., doi: 10.1038/s41551-025-01536-z (2025).

63. K. B. Baker, E. B. Plow, S. Nagel, A. B. Rosenfeldt, R. Gopalakrishnan, C. Clark, A. Wyant, M. Schroedel, J. Ozinga 4th, S. Davidson, O. Hogue, D. Floden, J. Chen, P. J. Ford, L. Sankary, X. Huang, D. A. Cunningham, F. P. DiFilippo, B. Hu, S. E. Jones, F. Bethoux, S. L. Wolf, J. Chae, A. G. Machado, Cerebellar deep brain stimulation for chronic post-stroke motor rehabilitation: a phase I trial. Nat. Med. 29, 2366–2374 (2023).

64. K. Kawahira, M. Shimodozono, S. Etoh, K. Kamada, T. Noma, N. Tanaka, Effects of intensive repetition of a new facilitation technique on motor functional recovery of the hemiplegic upper limb and hand. Brain Inj. 24, 1202–1213 (2010).

65. N. Bolognini, A. Pascual-Leone, F. Fregni, Using non-invasive brain stimulation to augment motor training-induced plasticity. J. Neuroeng. Rehabil. 6, 8 (2009).

66. K. K. Sellers, R. ’ee Gilron, J. Anso, K. H. Louie, P. R. Shirvalkar, E. F. Chang, S. J. Little, P. A. Starr, Analysis-rcs-data: Open-source toolbox for the ingestion, time-alignment, and visualization of sense and stimulation data from the Medtronic Summit RC+S system. Front. Hum. Neurosci. 15, 714256 (2021).

67. M. Olaru, S. Cernera, A. Hahn, T. A. Wozny, J. Anso, C. de Hemptinne, S. Little, W.-J. Neumann, R. Abbasi-Asl, P. A. Starr, Motor network gamma oscillations in chronic home recordings predict dyskinesia in Parkinson’s disease. Brain 147, 2038–2052 (2024).

68. N. V. Chawla, K. W. Bowyer, L. O. Hall, W. P. Kegelmeyer, SMOTE: Synthetic minority over-sampling technique. J. Artif. Intell. Res. 16, 321–357 (2002).

69. V. V. Moca, H. Bârzan, A. Nagy-Dăbâcan, R. C. Mureșan, Time-frequency super-resolution with superlets. Nat. Commun. 12, 337 (2021).

